# Aerosolized Hydrogen Peroxide Decontamination of N95 Respirators, with Fit-Testing and Viral Inactivation, Demonstrates Feasibility for Re-Use During the COVID-19 Pandemic

**DOI:** 10.1101/2020.04.17.20068577

**Authors:** T. Hans Derr, Melissa A. James, Chad V. Kuny, Devanshi Patel, Prem P. Kandel, Cassandra Field, Matthew D. Beckman, Kevin L. Hockett, Mark A. Bates, Troy C. Sutton, Moriah Szpara

**Affiliations:** Environmental Health and Safety and the Huck Institutes of the Life Sciences, Pennsylvania State University, University Park, Pennsylvania 16802, USA; Animal Resource Program, and the Huck Institutes of the Life Sciences, Pennsylvania State University, University Park, Pennsylvania 16802, USA; Occupational Medicine Program, and the Huck Institutes of the Life Sciences, Pennsylvania State University, University Park, Pennsylvania 16802, USA; Departments of Biology, Biochemistry and Molecular Biology, and the Huck Institutes of the Life Sciences, Pennsylvania State University, University Park, Pennsylvania 16802, USA; Plant Pathology and Environmental Microbiology, and the Huck Institutes of the Life Sciences, Pennsylvania State University, University Park, Pennsylvania 16802, USA; Veterinary and Biomedical Sciences, and the Huck Institutes of the Life Sciences, Pennsylvania State University, University Park, Pennsylvania 16802, USA; Statistics, and the Huck Institutes of the Life Sciences, Pennsylvania State University, University Park, Pennsylvania 16802, USA; Center for Infectious Disease Dynamics, and the Huck Institutes of the Life Sciences, Pennsylvania State University, University Park, Pennsylvania 16802, USA

**Keywords:** N95 respirators, filtering facepiece (FFP) respirators (FFR), decontamination, aerosolized hydrogen peroxide, COVID-19, SARS-CoV2, virologic testing, virus, fit-testing, disinfection, sterilization, CURIS^®^

## Abstract

In response to the demand for N95 respirators by healthcare workers during the COVID-19 pandemic, we evaluated decontamination of N95 respirators using an aerosolized hydrogen peroxide (aHP) system. This system is designed to dispense a consistent atomized spray of aerosolized, 7% hydrogen peroxide (H_2_O_2_) solution over a treatment cycle. Multiple N95 respirator models were subjected to ten or more cycles of respirator decontamination, with a select number periodically assessed for qualitative and quantitative fit testing. In parallel, we assessed the ability of aHP treatment to inactivate multiple viruses absorbed onto respirators, including phi6 bacteriophage, HSV-1, CVB3, and SARS-CoV-2. For pathogens transmitted via respiratory droplets and aerosols, it is critical to address respirator safety for reuse. This study provided experimental validation of an aHP treatment process that decontaminates the respirators while maintaining N95 function. External NIOSH certification verified respirator structural integrity and filtration efficiency after ten rounds of aHP treatment. Virus inactivation by aHP was comparable to the decontamination of commercial spore-based biological indicators. These data demonstrate that the aHP process is effective, with successful fit-testing of respirators after multiple aHP cycles, effective decontamination of multiple virus species including SARS-CoV- 2, successful decontamination of bacterial spores, and filtration efficiency maintained at or greater than 95%. While this study did not include extended or clinical use of N95 respirators between aHP cycles, these data provide proof of concept for aHP decontamination of N95 respirators before reuse in a crisis-capacity scenario.

**Importance:** The COVID-19 pandemic led to unprecedented pressure on healthcare and research facilities to provide personal protective equipment. The respiratory nature of the SARS-CoV2 pathogen makes respirator facepieces a critical protection measure to limit inhalation of this virus. While respirator facepieces were designed for single-use and disposal, the pandemic increased overall demand for N95 respirators, and corresponding manufacturing and supply chain limitations necessitated the safe reuse of respirators when necessary. In this study, we repurposed an aerosolized hydrogen peroxide (aHP) system that is regularly utilized to decontaminate materials in a biosafety level 3 (BSL3) facility, to develop methods for decontamination of N95 respirators. Results from virus inactivation, biological indicators, respirator fit testing, and filtration efficiency testing all indicated that the process was effective at rendering N95 respirators safe for reuse. This proof-of-concept study establishes baseline data for future testing of aHP in crisis capacity respirator-reuse scenarios.

## Introduction

The ongoing severe acute respiratory syndrome coronavirus 2 (SARS-CoV-2) pandemic resulted in a shortage of personal protective equipment (PPE). In healthcare settings, the need for PPE is critical to protect frontline healthcare providers from infection, and to reduce cross-contamination between patients with coronavirus disease 2019 (COVID-19) and other uninfected patients. In healthcare settings, N95 filtering facepiece (FFP) respirators, including surgical N95 respirators, are used to provide protection from airborne infectious particles. The N95 terminology refers to the ability to block at least 95 percent of the most penetrating particle sizes (0.1 - 0.3 micron). Proper use of N95 respirators requires qualitative fit-testing (QLFT) or quantitative fit-testing (QNFT), which are designed to ensure a tight face-to-respirator seal for each specific wearer’s facial characteristics.

The shortage of N95 respirators resulted from limitations on the required raw materials, limited capacity to manufacture respirators, and the ability of supply and distribution chains to handle increased global demand. For this reason, researchers sought to demonstrate the potential for decontamination and reuse of existing N95 respirators. Standardized procedures are well-established for the decontamination and reuse of medical equipment, such as autoclaving, steam treatment, and chemical inactivation (e.g. bleach) (1). Decontamination of most medical equipment is verified using spore-based biological indicators (2). Unlike most hospital equipment (e.g. steel, metal, plastic) or fabrics (e.g. blankets) for which standardized decontamination methods exist (1), N95 respirators are generally not intended for re-use (3).

Thus many of the standard decontamination approaches deform, damage, or destroy the integrity of N95 respirator fabric, nosepiece materials, or elastic straps (4–8). Hydrogen peroxide (H_2_O_2_)- based methods have been successfully adapted for use in decontamination of N95 respirators (4, 6–22), with indications that these methods are less damaging than other decontamination methods (e.g. chemical or steam) and can penetrate the densely-woven fabric of respirator facepieces (4, 6–8, 14, 22). In addition, the viricidal capability of H_2_O_2_ decontamination methods has been previously demonstrated (23–25). Vapor-phase H_2_O_2_ methods (e.g. “VHP” and other patented methods) have been used to decontaminate N95 respirators and were granted temporary U.S. Food and Drug Administration (FDA) “Emergency Use Authorization” (EUA) for healthcare use during the pandemic (26). However, these methods employ high concentrations of hydrogen peroxide (30-70%) and elevated temperature to achieve vaporization. Vapor-phase H_2_O_2_ (VHP) methods at these concentrations may pose increased health risks to decontamination personnel, and combined with elevated temperature, can result in notable respirator material decay (27). Historical aerosolized H_2_O_2_ (aHP) methods utilize lower peroxide concentrations (5- 6%) with silver ions and other antimicrobial agents, activated aerosolization via plasma, nozzle pressure, or ultrasound, with similar limitations to VHP methods (28–32). These methods have not received strong comparative support in U.S. markets due to lower decontamination effectiveness (30–38). Therefore, we utilized a recently-developed aHP method (i.e. the CURIS^®^ system), which dispenses a low concentration hydrogen peroxide solution through a precision adjustable nozzle (39). The unit design and decontamination process characteristics allow consistent distribution of disinfectant over time and enable effective decontamination of space and materials. The aHP approach has the potential to scale up for large clinical settings, since the number of respirators that can be decontaminated simultaneously is limited only by the room size.

At present, respirator manufacturers have not approved protocols for N95 respirator decontamination (3). To address imminent pandemic needs, healthcare settings have referenced U.S. Centers for Disease Control and Prevention (CDC) guidelines on provisional N95 respirator decontamination and reuse (40, 41). Battelle Memorial Institute of Columbus, Ohio, received FDA EUA status for an N95 decontamination protocol using a VHP method, based on a prior study addressing the potential for respirator reuse in emergency scenarios (4, 26). This study included evaluation of respirator structure, filtration, and manikin fit-testing, and used bacterial spore-based biological indicators to demonstrate effective decontamination (4); however, viral inactivation testing and respirator fit on live subjects were not performed. Since the current pandemic entails a respiratory viral pathogen, multiple decontamination protocols have been actively investigated at research universities and medical centers (6–21).

Given the reduced personnel health risks of using aHP, we assessed the ability of an aHP decontamination protocol to achieve viral and microbial decontamination of N95 respirators, while preserving respirator fit and integrity over multiple treatment cycles. Several respirator models in use by local healthcare and research personnel were included. Viral decontamination was tested using multiple species representing a range of pathogen characteristics: *Pseudomonas* phi6 bacteriophage (phi6), herpes simplex virus 1 (HSV-1), coxsackievirus B3 (CVB3), and SARS-CoV-2. Commercial *Geobacillus stearothermophilus* spore-based biological indicators were used in parallel throughout the aHP process to verify effectiveness of decontamination. We measured the inactivation of viruses by passive drying and by active aHP decontamination.

Fitness of respirators for re-use was assessed by qualitative and quantitative respirator fit-testing after decontamination, including up to 10 cycles of aHP treatment. Respirator structure and filtration efficiency testing was also performed. We also acquired real-time and diffusion sampler analyses of hydrogen peroxide levels throughout the decontamination process to monitor user safety.

## Results

This study was intended to rigorously evaluate a protocol for decontamination and re-use of N95 respirators using aerosolized hydrogen peroxide (aHP) treatment (7% H_2_O_2_; Curoxide^®^). The N95 respirator facepiece models examined here represent those frequently used at Penn State or within the Penn State Health system. Six N95 respirator models were selected, with the greatest number available being the 3M 8511 model (Table 1, Figure S1). The decontamination process was performed in the BSL3 enhanced facility, enabling the assessment of viral inactivation across multiple biosafety levels, including SARS-CoV-2 (Table 2, Figure S2). The BSL3 facility employs aHP on a routine basis to decontaminate solid equipment, and we adapted this protocol to account for the absorbent nature of N95 respirators. Standard aHP charge, pulse, and dwell period parameters were adjusted to optimize cycle times (Table 3) and account for biological indicator and virology results. Our final aHP process (matched to room size) utilized a 11:43 charge period to establish of aHP, followed by six pulse charges evenly spaced over 30 minutes to maintain H_2_O_2_ concentrations, and a 20-minute dwell period (Table 3).

**Table 1:** N-95 respirator facepiece models included in this study.

| Brand & model | # in study | Style | Type | Exhalation valve | Notes |
| --- | --- | --- | --- | --- | --- |
| 3M 8511 | 77 | molded | Non-surgical | yes |  |
| 3M 1860 | 10 | molded | Surgical | no |  |
| 3M 1870+ | 11 | folded | Surgical | no | Highest fluid resistance* |
| 3M 9211+ | 12 | folded | Non-surgical | yes | Same fabric as 1870+ |
| Honeywell Sperian N11125 | 5 | molded | Non-surgical | yes |  |
| Alpha ProTech | 65 | folded | Surgical | No |  |

\* Highest level of fluid resistance according to ASTM F1862 at 160 mm Hg (42).

**Table 2.** Characteristics of virus species used to test inactivation by aerosolized H2O2, as compared to SARS-CoV-2

| Virus species (abbreviation, taxonomic family) | Diameter | Capsid / virion shape | Genome type, ~size* | Titer inactivated by aHP^ | Biosafety Level (BSL) |
| --- | --- | --- | --- | --- | --- |
| Severe acute respiratory syndrome coronavirus 2 (SARS-CoV-2, *Coronaviridae*) | 120 nm | Enveloped, no icosahedral capsid | linear (+) ssRNA genome, ~30 kbp | $1.6 \times 10^5$ PFU/ml | BSL3 |
| Herpes simplex virus 1 (HSV-1, *Herpesviridae*) | 200 nm | Enveloped, icosahedral | linear dsDNA genome, ~152 kbp | $2.0 \times 10^6$ PFU/ml | BSL2 |
| Coxsackie virus B3 (CVB3, *Picornaviridae*) | 30 nm | Non-enveloped (naked), icosahedral | linear (+) ssRNA genome, ~7.4 kbp | $5.9 \times 10^4$ PFU/ml | BSL2 |
| *Pseudomonas* phi6 bacteriophage (phi6, *Cystoviridae*) | 85 nm | Enveloped, icosahedral | segmented, dsRNA genome, ~13.3 kbp | $2.4 \times 10^8$ PFU/ml | BSL1 |

\*ds, double-stranded; ss, single-stranded; ssRNA genomes are either (+) positive or (-) negative sense.
^Each viral species was tested for decontamination at the maximum available titer.

**Table 3:** Optimization of aHP treatment parameters

| **aHP**<br>**Cycle #** | **Room**<br>**Volume (ft<sup>3</sup>)** | **Charge**<br>**Period** | **Pulse**<br>**Period** | **Dwell**<br>**Period** | **aHP**<br>**Parameter Set** |
| --- | --- | --- | --- | --- | --- |
| 1a | 1700 | 16:20 | 40:00 | 0:00 | Initial |
| 1b | 1700 | 16:20 | 40:00 | 0:00 | Initial |
| 2 | 1700 | 11:43 | 30:00 | 0:00 | Modification 1 |
| 3 | 1700 | 11:43 | 30:00 | 0:00 | Modification 1 |
| 4 | 1700 | 11:43 | 30:00 | 0:00 | Modification 1 |
| 5 | 1700 | 11:43 | 30:00 | 20:00 | Final |
| 6 | 1700 | 11:43 | 30:00 | 20:00 | Final |
| 7 | 1700 | 11:43 | 30:00 | 20:00 | Final |
| 8 | 1700 | 11:43 | 30:00 | 20:00 | Final |
| 9 | 1700 | 11:43 | 30:00 | 20:00 | Final |
| 10 | 1700 | 11:43 | 30:00 | 20:00 | Final |
| 11 | 1700 | 11:43 | 30:00 | 20:00 | Final |
| 12 | 1700 | 11:43 | 30:00 | 20:00 | Final |
| Post 1 | 1840 | 12:41 | 30:00 | 20:00 | Final* |
| Post 2 | 1840 | 12:41 | 30:00 | 20:00 | Final* |
| Post 3 | 1840 | 12:41 | 30:00 | 20:00 | Final* |

\* These cycles were carried out in a larger room, which necessitated adjustment to the charge time to account for the larger room volume.

### Chemical Verification and Biological Validation of Aerosolized H2O2 Process

Chemical and bacterial spore-based biological indicators (BIs) were used to verify aHP treatment and decontamination. All chemical indicators located throughout the treatment area (Prep Room) during all aHP cycles confirmed exposure to hydrogen peroxide. For cycles in which charge, pulse, and dwell periods were utilized, all biological indicators (BIs) similarly indicated successful decontamination, with the exception of aHP cycles 3 and 5 (Table 4, Figure S3). In cycle 3 the additional dwell period was not yet implemented, and BIs indicated an unsuccessful decontamination cycle (1 positive, 5 negative). External contamination of one spore coupon after treatment cycle 5 (via dropping) likely resulted in the single positive indicator for this cycle (Table 4). Overall chemical and biological indicator results indicated successful aHP treatment and decontamination of N95 respirators.

**Table 4:** Spore-based Biological indicator (BI) and Virus Inactivation Results

| aHP Cycle # | aHP-treated Spore-based BIs | Control Spore-based BIs | Spore-based BI Results | aHP parameter set | Viruses Tested* | Virus Inactivation Results |
| --- | --- | --- | --- | --- | --- | --- |
| 1a | 12 | 1 | PASS | Initial |  |  |
| 1b | 12 | 1 | PASS | Initial |  |  |
| 2 | -- | -- | -- | Modification 1 |  |  |
| 3 | 6 | 1 | FAIL (1+/5-) | Modification 1 | phi6, HSV1, CVB3 | 3 of 64 sites positive |
| 4 | 6 | 1 | PASS | Modification 1 |  |  |
| 5 | 6 | 1 | FAIL^ (1+/5-) | Final |  |  |
| 6 | 6 | 1 | PASS | Final | HSV1, CVB3 | 1 of 26 sites positive |
| 7 | 6 | 1 | PASS | Final |  |  |
| 8 | 6 | 1 | PASS | Final |  |  |
| 9 | 6 | 1 | PASS | Final |  |  |
| 10 | 6 | 1 | PASS | Final |  |  |
| 11 | 6 | 1 | PASS | Final |  |  |
| 12 | 6 | 1 | PASS | Final |  |  |
| Post 1 | -- | -- | -- | Final | phi6, HSV1, CVB3 | 0 of 66 sites positive |
| Post 2 | 12 | 1 | PASS | Final | SARS-CoV-2 | 0 of 24 sites positive |
| Post 3 | 12 | 1 | PASS | Final | SARS-CoV-2 | 0 of 24 sites positive |

\* The N95 respirators used for virus inoculation at each cycle were subjected to all preceding aHP cycles – e.g. respirators used for viral testing in aHP cycle #6 had been through 5 prior aHP cycles. For cycles labeled “Post”, the respirators for viral testing had been through 10-12 prior aHP cycles.
^A spore disc dropped during transfer to media was the suspected cause of this failure.

### Real-Time Hydrogen Peroxide Monitoring

Real-time measurement of H_2_O_2_ concentrations during decontamination were obtained with a portable, real-time ATI PortaSens II monitor (Table 5). H_2_O_2_ concentrations in the treatment area were measured at ≥ 120 ppm during the charge and pulse periods (maximum sensor capability).

**Table 5:** Portable real-time H2O2 measurements, using ATI PortaSens II

| aHP Cycle ID | Measurement Location* | Measurement Condition | H<sub>2</sub>O<sub>2</sub> Concentration or range (ppm)** | Time period to decayed concentration (minutes) | aHP Parameter set |
| --- | --- | --- | --- | --- | --- |
| 1a | IC, wall port | Charge | > 120 | -- | Initial |
| 1a | IC, wall port | Aeration, to entry | 2.5 – 57 | 52 | Initial |
| 1b | IC, wall port | Charge | > 120 | -- | Initial |
| 2 | No data | No data | No data | No data | Modification 1 |
| 3 | IC, door open | Aeration, to entry | 1.5 – 11.6 | 20 | Modification 1 |
| 3 | IC, door open | Aeration, to respirator measurement | 1.5 – 0.8 | 40 | Modification 1 |
| 4 | IC, door open | Aeration, to entry | 2 – 10.6 | 20 | Modification 1 |
| 4 | respirator surface | After overnight drying | 0 – 3 | overnight | Modification 1 |
| 5 | IC, wall port | Charge | > 120 | -- | Final |
| 6 | IC, wall port | Pulse | 120 | -- | Final |
| 6 | respirator surface | After overnight drying | 0 | overnight | Final |
| 7 | IC, wall port | Charge | > 120 | -- | Final |
| 7 | OC, seal check(s) | Charge, Pulse, Dwell | 0 | -- | Final |
| 7 | IC, door open | Aeration, to entry | 2 – 7 | 27 | Final |
| 7 | respirator surface | After overnight drying | 0 | overnight | Final |
| 8 | IC, wall port | Pulse | 88 | -- | Final |
| 9 | IC, wall port | Charge | > 120 | -- | Final |
| 9 | IC, door open | Aeration, to entry | 1.6 – 24 | 20 | Final |
| 9 | respirator surface | Aeration, to respirator measurement | < 1 | 85 | Final |
| 10 | OC, seal check | Pulse, door seal repair | 0 – 1.6 | -- | Final |
| 10 | IC, door open | Aeration, to entry | 1.3 – 15 | 32 | Final |
| 10 | respirator surface | After overnight drying | < 1 | overnight | Final |
| 11 | IC, door open | Aeration, to entry | 1 – 13.6 | 35 | Final |
| 11 | respirator surface | After overnight drying | < 1 | overnight | Final |
| Post | respirator surface | Aeration to 1ppm, respirator data log | 1 – 8.1 | 150 | Final |

\* Measurement Location: IC – inside containment / treatment area, (wall port) – through sealable wall port, (door open) – door ajar after door seal removed, OC – outside containment / treatment area, (seal check) – Prep Room door tape seal, (respirator surface) – at respirator surface following room entry.
\*\* Low measurement value represents lowest concentration following concentration decay; high value represents starting concentration (prior to decay).

Hydrogen peroxide concentrations outside the sealed entryway were low or undetected (0 ppm). On occasions where a broken seal arose in the tape around the door (Figure S2), low concentrations were detectable outside the door (up to 3 ppm). Once corrected, these levels immediately dropped to 0 ppm. During aeration after aHP treatment, H_2_O_2_ levels were monitored until concentrations measured ≤ 2 ppm. At this time research personnel entered the Prep Room to measure H_2_O_2_ concentrations at respirator surfaces. Hydrogen peroxide concentrations were observed to decline rapidly during aeration within 20-30 minutes of door opening (Table 5).

Initial respirator surface concentrations (at fabric) typically exceeded 2 ppm. Subsequent respirator drying to achieve < 1 ppm required further room aeration (exhaust ventilation) exceeding two hours, or overnight (Table 5). Once H_2_O_2_ respirator surface concentrations measured < 1 ppm, they were either subjected to further cycles of aHP, or collected for fit-testing or virus inactivation analysis.

### Qualitative and Quantitative Fit-Testing After aHP Decontamination

We employed both qualitative (QLFT) and quantitative (QNFT) fit-testing. QLFT allowed repeated non-destructive testing across multiple rounds of decontamination, while QNFT provided numerical data on fit factor and ruled out any false negatives due to user fatigue. QLFT and QNFT was conducted after the first, fifth, and tenth decontamination cycles for both a male and female subject. These tests included a large number of 3M model 8511 respirators (for which we had the most available in the starting pool), and a smaller number of 5 other respirator models (Table 1, Figure 1). All respirators passed all QLFT and QNFT fit tests (Figure 1). One respirator facepiece (3M model 1870+) experienced a broken strap after the eighth cycle of aHP; this respirator had passed QLFT successfully in earlier cycles but was excluded from final QNFT. No other failures occurred, therefore with 95% confidence, at least 95% of 3M model 8511 respirators maintained successful fit seal after ten aHP decontamination cycles. QNFT results with 3M model 8511 respirators passed the minimum passing fit factor of 100, and the maximum quantifiable fit factor of 200(+) in 8 of 9 tests. The numerical range of passing fit factors (100–200), and available sample quantity, limit any further statistical analysis. The successful fit-testing results indicate that repeated cycles of aHP decontamination do not interfere with respirator fit for reuse.

**Figure 1:**
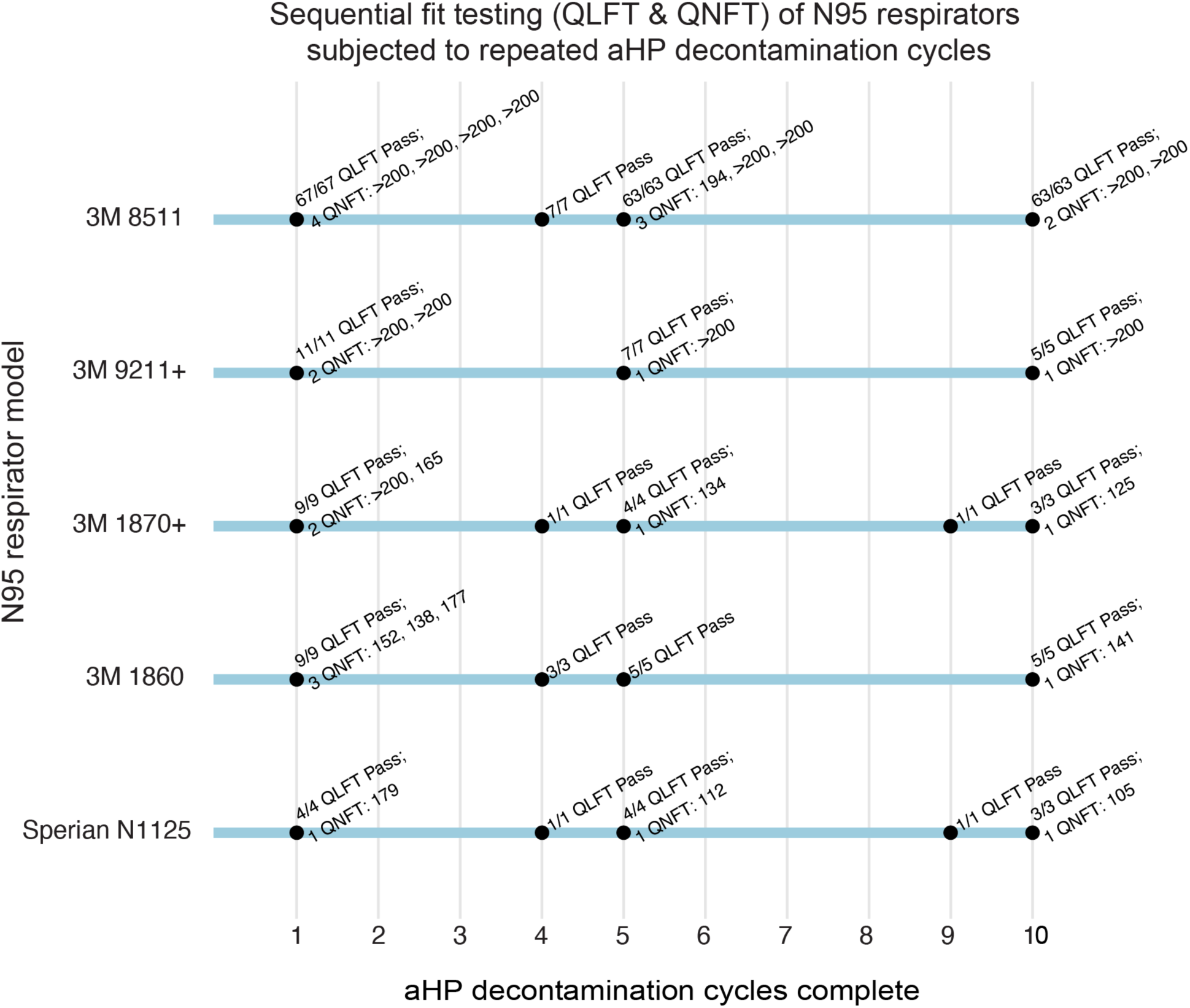
Sequential fit testing (QLFT & QNFT) of N95 respirators subjected to repeated aHP decontamination cycles. Results demonstrate that all 3M model 8511 respirators successfully pass QLFT and QNFT after 1, 5, and 10 cycles. In particular, 8 of 9 QNFT results for the 3M model 8511 respirator surpassed a fit factor of 200, the maximum reportable by the test method – providing qualitative, yet objective, evidence of the safety margin related to fit integrity after 10 aHP decontamination cycles. All respirator models except Alpha Protech (which had inconsistent fit-test results; see Methods for details) passed all QLFT and QNFT to which they were subjected. Models Alpha ProTech (aHP cycle 1) and Kimberly Clark (aHP cycle 2) were only included for a single cycle and are thus not shown here.

### Respirator Filtration Efficiency Testing

N95 respirator filtration efficiency testing was conducted on aHP-treated 3M 8511 respirators using the NIOSH test protocol under full-load test conditions (Table 6). No visible degradation was found on inspection (e.g. metal nose guard discoloration, unusual thinning, or wear).

**Table 6:**
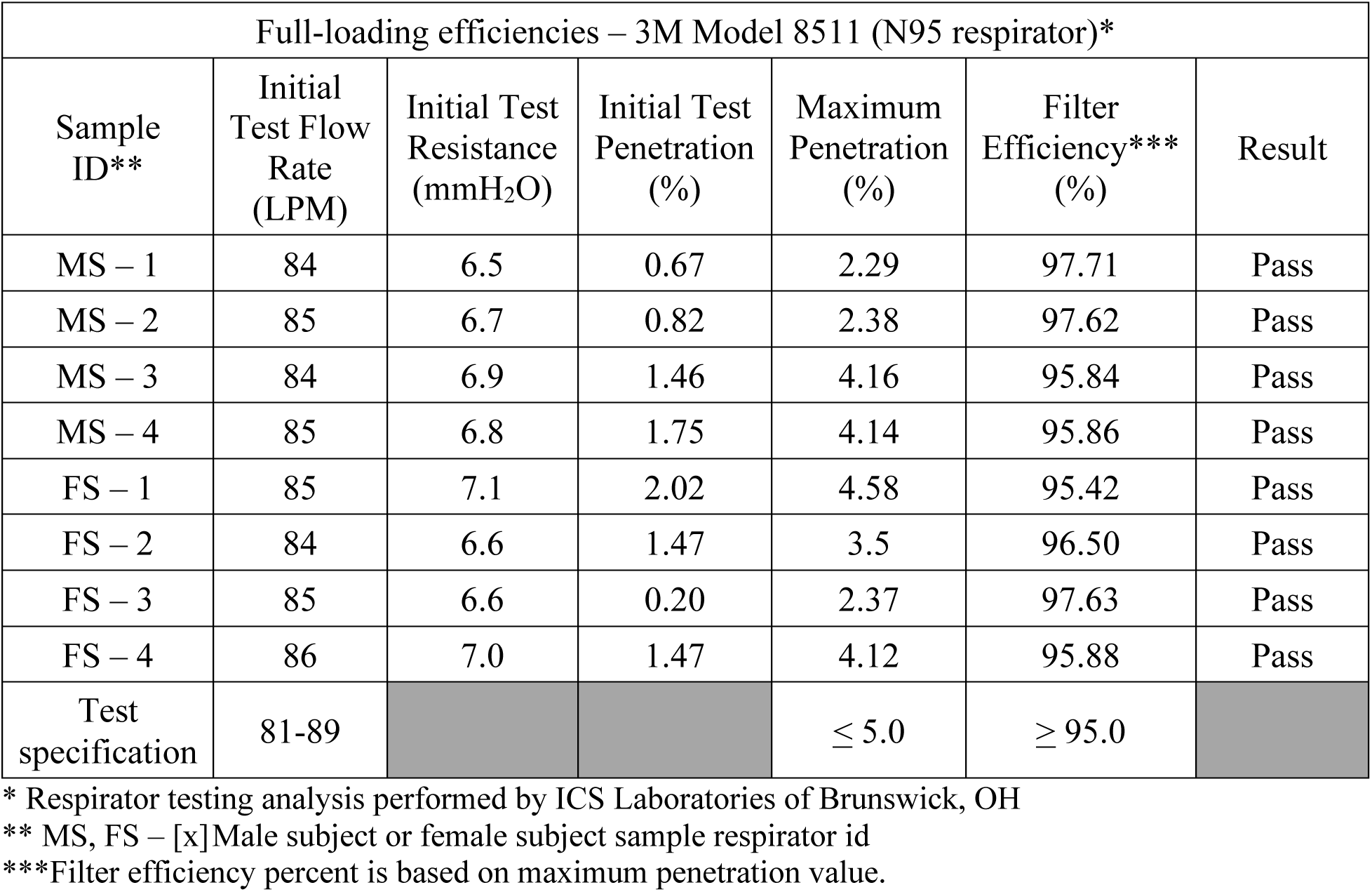
Respirator filtration efficiency testing results following 10 cycles of aerosolized hydrogen peroxide decontamination

| Full-loading efficiencies – 3M Model 8511 (N95 respirator)* |  |  |  |  |  |  |
| --- | --- | --- | --- | --- | --- | --- |
| Sample ID** | Initial Test Flow Rate (LPM) | Initial Test Resistance (mmH<sub>2</sub>O) | Initial Test Penetration (%) | Maximum Penetration (%) | Filter Efficiency*** (%) | Result |
| MS – 1 | 84 | 6.5 | 0.67 | 2.29 | 97.71 | Pass |
| MS – 2 | 85 | 6.7 | 0.82 | 2.38 | 97.62 | Pass |
| MS – 3 | 84 | 6.9 | 1.46 | 4.16 | 95.84 | Pass |
| MS – 4 | 85 | 6.8 | 1.75 | 4.14 | 95.86 | Pass |
| FS – 1 | 85 | 7.1 | 2.02 | 4.58 | 95.42 | Pass |
| FS – 2 | 84 | 6.6 | 1.47 | 3.5 | 96.50 | Pass |
| FS – 3 | 85 | 6.6 | 0.20 | 2.37 | 97.63 | Pass |
| FS – 4 | 86 | 7.0 | 1.47 | 4.12 | 95.88 | Pass |
| Test specification | 81-89 |  |  | ≤ 5.0 | ≥ 95.0 |  |

\* Respirator testing analysis performed by ICS Laboratories of Brunswick, OH
\*\* MS, FS – [x] Male subject or female subject sample respirator id
\*\*\*Filter efficiency percent is based on maximum penetration value.

Filtration efficiencies were found to exceed 95% for all eight respirators subjected to ten cycles of aHP decontamination. Therefore, for at least ten aHP cycles, there were no adverse impacts or loss of N95 respirator efficiencies. Two additional unused, untreated 3M 8511 respirators were additionally tested and found with slightly reduced filtration efficiencies (94.4%, 94.6%), consistent with the respirator manufacturer’s 5-year shelf-life limitation. These data indicate that overall respirator performance was maintained over time and after aHP treatment.

### Application of Multiple Viral Species to N95 Respirator Facepieces

Viral inactivation on respirator facepieces was anticipated to occur both by the passive process of drying or desiccation, and by the active process of aHP decontamination. Multiple virus species were included to test the decontamination potential of aHP against viruses in general, as well as against SARS-CoV-2. These included phi6, HSV-1, CVB3, and SARS-CoV-2 (see Table 2 and Methods for details). Respirators used for virus inactivation testing were those previously subjected to aHP treatment and fit-testing, to spare overall respirator consumption (see Table 4 and Methods). Initial application of virus to different respirator facepiece types revealed clear differences in relative absorption vs. fluid repulsion (Figure S1). 3M respirator models 1870+ and 9211+ share a common outer fabric which is listed by the manufacturer as having the highest fluid resistance of any N95 respirator (Table 1) (42). In our testing, these two respirator models displayed no apparent absorption of virus inoculum and instead dried with a “coffee ring effect”. All other respirator types (Table 1) experienced a combination of liquid spreading, absorption, and evaporative drying of the virus inoculum droplet. Viruses were inoculated onto different areas of each respirator facepiece model, including the outer and inner fabric surfaces, the elastic strap, and where present, the inner and outer surface of the plastic exhalation valves.

### Decontamination of Virus-Inoculated Respirators by Aerosolized H2O2 Treatment

We set out to assess the effectiveness of aHP treatment for active decontamination of virus inoculated onto N95 respirators. Virus-inoculated respirators were subjected to aHP treatment, using both “Modification 1” and “Final” parameters (Table 3). Viral testing of decontamination was conducted during five independent aHP cycles (Table 4), using the maximum inoculum titer available for each viral stock preparation. For phi6 bacteriophage, this included 34 “aHP-treated” sites spanning two independent rounds of testing (Figure 2). For HSV-1 and CVB3, this included 62 and 60 “aHP-treated” sites respectively, spanning three independent rounds of testing (Figure 3 and Figure 4). Across a total of 204 respirator sites inoculated with one of four virus species tested (Table 2; see also Table S1), only four sites had any detectable virus remaining. Three of these rare positive virus plaques were detected in an aHP cycle using the “Modification 1” parameters (Figure 2A and 3A; Table 4; see also Figure S4) – these were a key motivation to add the dwell time for the “Final” aHP cycle parameters (Table 3). Overall, aHP treatment produced a 4-7 log_10_ reduction in viral load (10^7^ reduction for phi6, Figure 2A; 10^5^ reduction for HSV-1, Figure 3; 10^4^ reduction for CVB3; Figure 4). There was no observable difference in the effectiveness of aHP decontamination for inner vs. outer surfaces of respirators (Figures 2-5; see also Table S1), or in limited testing of alternative inoculation sites such as elastic straps (Figure 3, Figure 4, Table S1). The success of virus inactivation by aHP treatment mirrored the results of spore-based biological indicators (Table 4).

**Figure 2:**
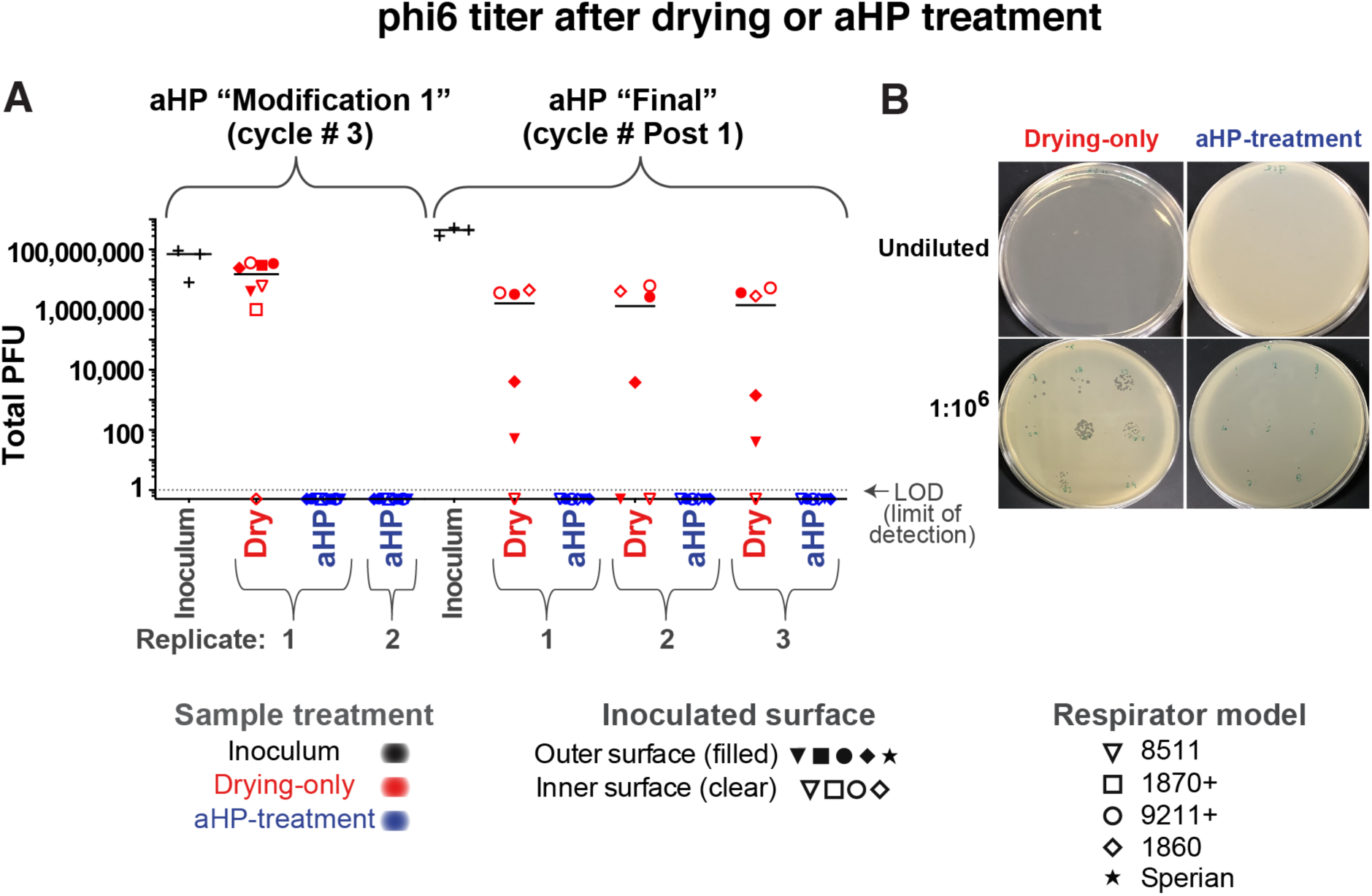
Infectious titer of phi6 bacteriophage inoculated on N95 respirator facepieces was eliminated after aerosolized H_2_O_2_ (aHP) decontamination. (A) Data are plotted for each aHP cycle in which viral testing was done (see Table 4). Multiple models of N95 respirator (see Table 1) were inoculated and either treated as “drying only” controls (red) or subjected to aHP treatment (blue). The respirator surface and model are indicated by the symbol shape and fill. The median of all points within a given aHP cycle and treatment is indicated by a solid horizontal line. The dashed horizontal line indicates the limit of detection (LOD) at 1 viral plaque-forming unit (PFU), in the resuspended but undiluted volume from the site of viral inoculation. (B) Image depicts Petri dish plating of bacterial lawns exposed to phi6 from drying-only (left side) or aHP-treated (right side) respirator inoculation sites. These were applied to the bacterial lawn either as an undiluted resuspension (top row) or 1:10^6^ dilution applied to focal points (bottom row). For the purposes of illustrating the decontaminated sites where zero plaques were detected, these numbers were replaced with fractional values (0.5), to allow their visualization on this log-scale plot. See Table S1 for all data values.

**Figure 3:**
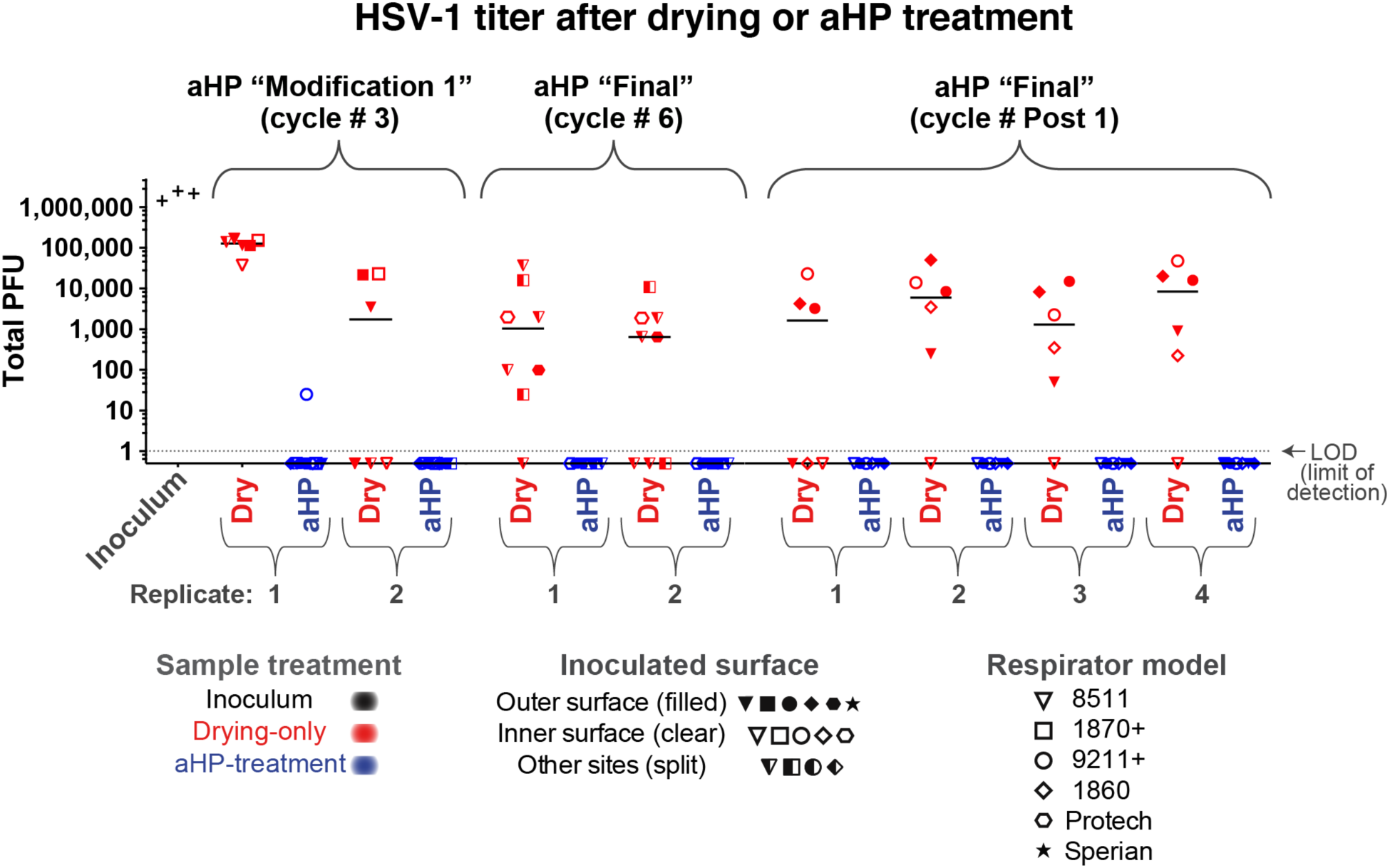
Infectious titer of HSV-1 inoculated on N95 respirator facepieces was reduced by drying and eliminated after aerosolized H_2_O_2_ (aHP) decontamination. Data are plotted for each aHP cycle in which viral testing was done (see Table 4). For HSV-1, the sole positive plaque after aHP treatment occurred in aHP cycle #3, when the “Modification 1” parameters were in use (Tables 3-4). This failure, in concert with a spore-based biological indicator and 2 CVB3 plaques (Figure 4) motivated the addition of a “dwell time” in the final aHP parameters. As in Figure 2, multiple models of N95 respirator (see Table 1) were inoculated and either treated as “drying only” controls (red) or subjected to aHP treatment (blue). The respirator surface and model are indicated by the symbol shape and fill. The median of all points within a given aHP cycle and treatment is indicated by a solid horizontal line. The dashed horizontal line indicates the limit of detection (LOD) at 1 viral plaque-forming unit (PFU), in the resuspended but undiluted volume from the site of viral inoculation. For the purposes of illustrating the decontaminated sites where zero plaques were detected, these numbers were replaced with fractional values (0.5), to allow their visualization on this log-scale plot. See Table S1 for all data values.

**Figure 4:**
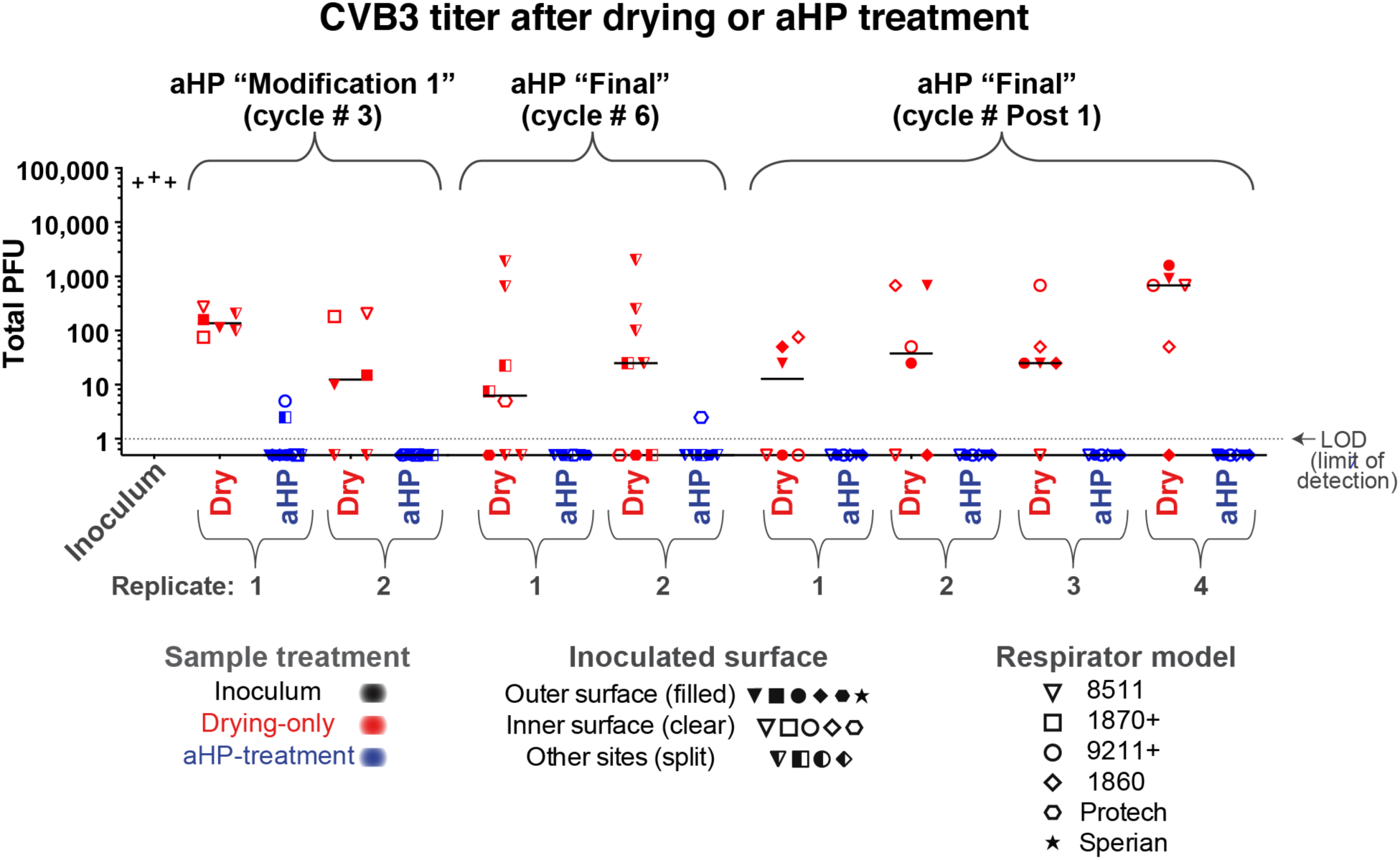
Infectious titer of CVB3 inoculated on N95 respirator facepieces was reduced by drying and eliminated after aerosolized H_2_O_2_ (aHP) decontamination. Data are plotted for each aHP cycle in which viral testing was done (see Table 4). For CVB3, two positive plaques after aHP treatment occurred in aHP cycle #3, when the “Modification 1” parameters were in use (Tables 3-4). This failure, in concert with a spore-based biological indicator and 1 HSV-1 plaques (Figure 4) motivated the addition of a “dwell time” in the final aHP parameter. The only other positive CVB3 plaque after aHP treatment occurred in aHP cycle 6 (B), and no plaques were detected in the replicate or in parallel samples. As in Figure 2, multiple models of N95 respirator (see Table 1) were inoculated and either treated as “drying only” controls (red) or subjected to aHP treatment (blue). The respirator surface and model are indicated by the symbol shape and fill. The median of all points within a given aHP cycle and treatment is indicated by a solid horizontal line. The dashed horizontal line indicates the limit of detection (LOD) at 1 viral plaque-forming unit (PFU), in the resuspended but undiluted volume from the site of viral inoculation. For the purposes of illustrating the decontaminated sites where zero plaques were detected, these numbers were replaced with fractional values (0.5), to allow their visualization on this log-scale plot. See Table S1 for all data values.

**Figure 5:**
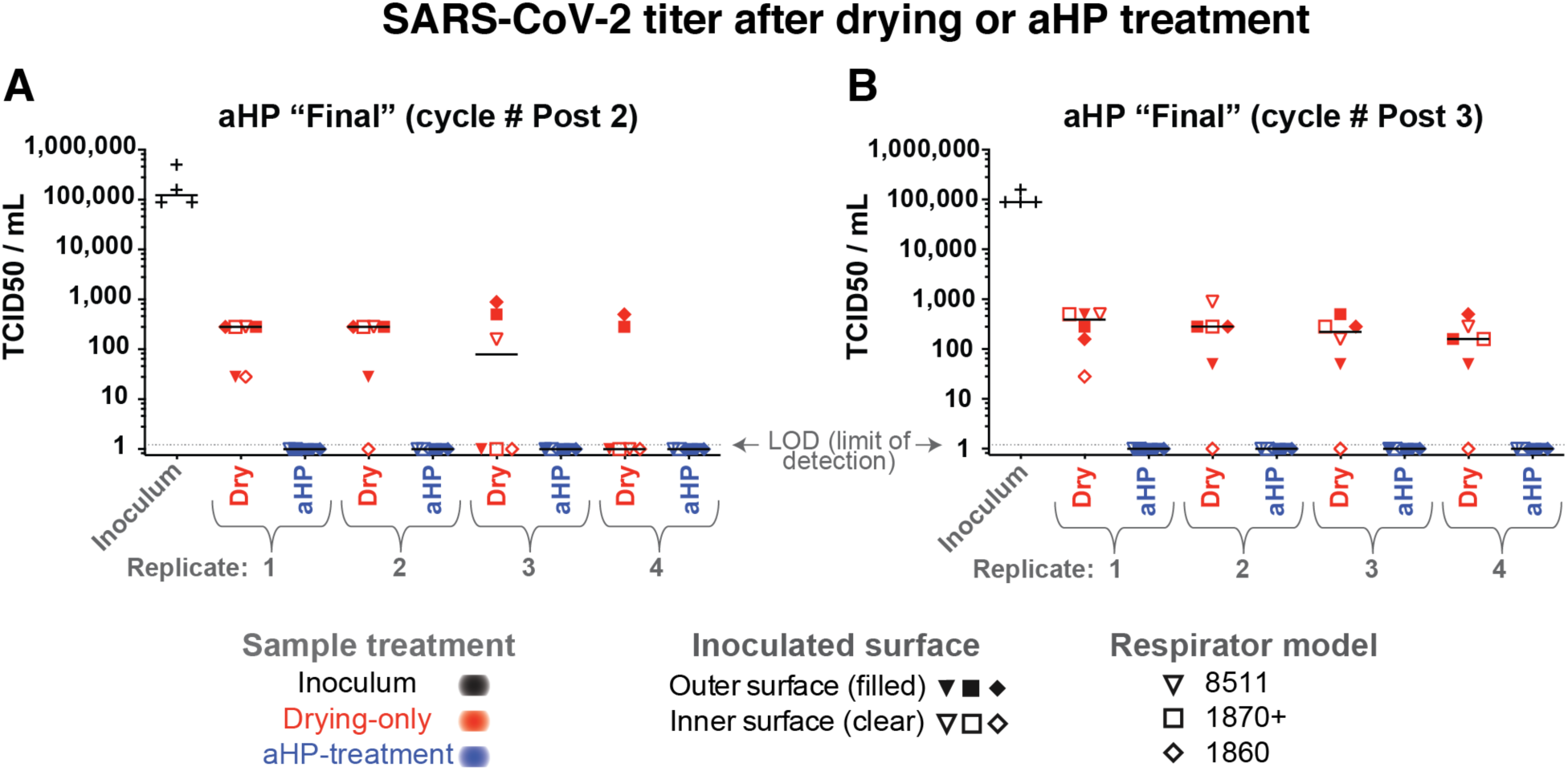
Infectious titer of SARS-CoV-2 inoculated on N95 respirator facepieces was reduced by drying and eliminated after aerosolized H2O2 (aHP) decontamination. (A-B) Data are plotted separately for each aHP cycle in which viral testing was done (see Table 4). For SARS-CoV-2, no infectious virus was detected by TCID50 assay after aHP treatment. As in Figure 2, multiple models of N95 respirator (see Table 1) were inoculated and either treated as “drying only” controls (red) or subjected to aHP treatment (blue). The respirator surface and model are indicated by the symbol shape and fill. The median of all points within a given aHP cycle and treatment is indicated by a solid horizontal line. Viral titer was determined by tissue culture infectious dose 50 (TCID50) assay in 96-well plates, with a limit of detection (LOD) of 1.2 (see Methods for details). For the purposes of illustrating the decontaminated samples where no virus was detected, these numbers were plotted as a value of 1. See Table S1 for all data values.

### Inactivation of Multiple Viral Species by Air-Drying on Respirators

Since passive viral inactivation by drying likely occurs during the time involved in decontamination, we measured the amount of virus remaining on inoculated but untreated N95 respirators. For each cycle of viral testing, the duration of passive viral inactivation was matched to the duration of the decontamination process (including aHP treatment, subsequent aeration, and transport). These “drying-only” samples confirmed a partial loss of viral infectiousness, ranging from 10-100-fold for phi6 and HSV-1, and 100-fold or greater for CVB3 and SARS-CoV-2 (Figures 2-5; see also Figure S4 and Table S1). For phi6 bacteriophage, this included 26 “drying-only” inoculation sites spanning three independent rounds of testing (Figure 2). For HSV-1 and CVB3, this included 52 “drying-only” inoculation sites for each virus species, spanning four independent rounds of testing (Figure 3 and Figure 4, respectively). Viral inactivation by drying was not markedly different on inner vs. outer surfaces of the respirator models (Figures 2-5; see also Table S1), or in limited testing of alternative inoculation sites, such as elastic straps or plastic exhalation valves (Figure 3A-B, Figure 4A-B, Table S1).

### Inactivation of SARS-CoV-2 by aHP Treatment

Experimental testing of SARS-CoV-2 by aHP necessitated that all work be completed at BSL3. These studies were conducted with the 3M 8511, 1860, and 1870+ N95 respirators and included two independent rounds of testing (Figure 5). As with the surrogate virus species, SARS-CoV-2 was first inoculated onto respirator facepieces. The “drying only” controls were left in the BSL3 ambient environment, while matched samples were subjected to aHP-treatment. The respirators used for SARS-CoV-2 testing had been subjected to 10 prior rounds of aHP before virus inoculation (Table 4). For SARS-CoV-2, viral testing included 48 “drying-only” inoculation sites and 48 “aHP-treated” sites, spanning two independent rounds of testing (Figure 5). We observed a partial loss of viral infectiousness for SARS-CoV-2 due to drying (e.g. 10^6.125^ TCID50/mL input vs ∼10^2^ TCID50/mL after drying; Figure 5). Importantly, no infectious SARS-CoV-2 remained on any respirator model after aHP decontamination (Figure 5).

## Discussion

Based on a series of ten respirator decontamination cycles and multiple rounds of viral inactivation testing, multiple N95 respirator models tested were found to be suitable for aHP decontamination and reuse. We found that respirators successfully passed qualitative respirator fit testing after multiple cycles of the aHP decontamination process, and ultimately passed tests indicating no loss in filtration efficiency. Most studies thus far have focused either on verifying fit-testing after respirator decontamination (6–10, 12, 15, 16, 20), or examined how decontamination approaches inactivate one or more virus species on respirators (6–8, 11, 13, 14, 18–21). A handful of N95 respirator decontamination studies have combined fit-testing with measures of viral inactivation (6–8), but none incorporated viral- or fit-testing after 5 or 10 decontamination cycles as done here, or included the parallel use of biological indicators. Only rarely have studies included extended-use between multiple rounds of decontamination cycles (10), although this is a key aspect of reuse that warrants further study. Our study is unique in including multiple measures of respirator integrity via fit-testing, as well as filtration efficiency, and robust verification of viral inactivation using BIs, multiple surrogate viruses, and SARS-CoV-2 in parallel (22). The breadth of this study aims to extend its usefulness beyond the current pandemic.

### Respirator Resilience for Re-Use

Aerosolized H_2_O_2_ decontamination of the N95 respirators used in this study did not indicate any adverse impact on final respirator filtration efficiency, although extended-use of respirators between aHP cycles was not possible in this study. We documented effective control of H_2_O_2_ levels, supporting effective pathogen decontamination with no detectable researcher exposure. These data support and reflect those of parallel studies that have tested respirator fit after decontamination by other forms of hydrogen peroxide vapor (VHP) and/or similar methods (6–10, 12, 15, 16, 20). Further studies will be needed to assess the H_2_O_2_ concentration profile inside containment, using real-time instruments with a wider detection range and automated data logging.

As noted above, most studies that include successful fit-testing and verification of viral inactivation have not pursued this testing across multiple (i.e. 10) cycles of decontamination (6–8, 20). Lab-based conditions such as those used here do not fully reflect clinical use conditions. While respirators were physically stretched between each decontamination cycle as a proxy for donning and doffing, sustained clinical use includes other stressors which may influence fit and performance (e.g. exhaled moisture or perspiration) (10, 16). However as noted by the CDC, respirators with obvious signs of use (e.g. makeup or patient fluids) should be discarded and not used for decontamination (40, 41). Other studies have explored respirator decontamination in these real-world use scenarios, albeit without including the parallel testing of viral species, BIs, and cycle numbers that were included here (10, 15–17, 20). We anticipate that future studies will address the implementation of aHP-based decontamination in a clinical-use setting.

During this study, one 3M 1870+ respirator suffered a broken rubber strap after eight cycles of aHP, during inter-cycle strap stretching. The breakage occurred at a point on the strap that corresponded with a penned hash mark (used to denote decontamination cycle; see Figure S2). There were no other instances of failure for this or any other respirator models. Prior groups have likewise noted strap-based failures after multiple cycles of respirator decontamination (4, 6, 19, 20). We recommend using care when marking respirators during re-use protocols.

### Viral and Biological Indicator Inactivation

Viral inactivation by drying depends on multiple factors including surface type, humidity, temperature, virion size and type, and duration of drying (43–46). All studies of N95 respirator decontamination include both the passive inactivation of viruses by air drying and their active decontamination by aHP or comparable treatment (43–46). Clinical exposure of respirators to SARS-CoV-2 or other viral pathogens would likewise entail ambient drying prior to any respirator decontamination or re-use (e.g. virus may dry onto an N95 in the course of a work-shift, or during bagging for decontamination or direct re-use). The “drying-only” time frame used here was shorter than used in studies modeling re-use in a clinical setting (10, 15–17, 20). This suggests that crisis-capacity protocols that involve respirator re-use after multiple days of drying, even without aHP or active decontamination, likely provide substantial levels of viral inactivation (40, 41).

To model diverse routes of respirator exposure to viral pathogens during use, we inoculated viruses onto different N95 models and surfaces to thoroughly test for the ability of aHP to inactivate viruses. This included different regions of the respirator (e.g. outer vs. inner surface) to discern whether any differences in these fabrics would influence viral inactivation. This work was inspired by early efforts to verify viral inactivation in the context of N95 respirator surfaces, such as that of Kenney et al (13). We found scant evidence of viral survival during the aHP decontamination process. Relative to studies using different decontamination methods to test viral inactivation on N95 respirators, these data show equivalent or better inactivation of viruses (6–8, 11, 13, 14, 18–21, 43).

Commercial bioindicator tests have been relied upon for verification of decontamination for decades (2, 47). Overall we observed parallel outcomes in terms of successful decontamination of viral species and bacterial spore-based BIs (Table 4), echoing the few other studies that have used these approaches in parallel (14, 18, 20). During establishment of aHP cycle parameters we noted a concomitant failure of BIs and viral inactivation in cycle 3, before the dwell period was added (Tables 3 and 4). The parallels in success or failure of both BIs and viral inactivation suggest that commercial spore-based BIs provide a useful predictor of success or failure for decontamination of N95 respirators, particularly in settings where direct viral testing is not feasible (2, 10, 15–17). The ten cycles of aHP decontamination achieved here are well beyond the CDC’s crisis-capacity plans, which recommend no more than five total rounds of respirator reuse (40, 41). While clinical use of respirators, by a multitude of healthcare workers, was not included in the present study, we foresee that such testing will be an important next step.

We used multiple virus species to test the viral inactivation capabilities of aHP decontamination. These viruses represented multiple characteristics of human viral pathogens, with a range of virion and genome types and sizes, and previously documented environmental stability (44, 45, 48). Like SARS-CoV-1, SARS-CoV-2 has a high level of environmental stability (49–53).

Coronaviruses have a lipid-enveloped virion of ∼120 nm, with no icosahedral capsid core, containing a single-stranded, linear, positive-sense RNA genome (Table 2). Virions of HSV-1 and phi6 bacteriophage have a lipid envelope, with an underlying icosahedral capsid core. In contrast, CVB3 has a non-enveloped or naked icosahedral capsid virus. Prior work has shown that naked-capsid viruses have a higher stability than enveloped viruses, thus demonstrating the range of aHP decontamination abilities (44, 45, 48). While most pathogens utilized here require propagation in mammalian cell lines at biosafety level 2 (BSL2), phi6 can be assayed more flexibly, using rapid bacterial cultures (24-hour turnaround) at biosafety level 1. Phi6 is a natural pathogen of the bacterial species *Pseudomonas syringae pathovar phaseolicola*, which is itself a pathogen of green beans. All viral species examined in this study were effectively decontaminated by aHP (Table 4). While the aerosolization of viruses was not incorporated here, this approach merits inclusion in future studies. Together, the combination of respirator fit testing and virus inactivation testing used here indicate that aHP is a viable decontamination process to enable crisis-capacity reuse of N95 respirators during viral pandemics.

## Methods

### Decontamination Facility

The decontamination process was carried out in the Eva J. Pell Laboratory for Advanced Biological Research at The Pennsylvania State University, University Park campus. This facility is a purpose-built BSL3 enhanced facility, and all required approvals were obtained from the Institutional Biosafety Committee (IBC) for work involving viruses, as described below.

The primary decontamination process was performed within in an approximate 1,700 cubic feet sealed Preparation Room (Prep Room), followed by additional virus inactivation testing with SARS-CoV-2 in a separate, nearby 1,840 cubic feet Prep Room. Procedures and personal protective equipment (PPE) suitable for the viruses and materials in use were strictly observed in this biosafety level 3 (BSL-3) facility, which has consistently maintained institutional, CDC, and USDA approval for work with risk group 3 pathogens since its commissioning in 2014.

### Decontamination Preparation

Respirators for decontamination were staged on a portable metal rack located centrally in the Prep Room (Figure S2). Filtered and conditioned air is supplied to the Prep Room and the air exhausted from the room is HEPA-filtered. Bubble-tight dampers (Camfil Farr^®^) were operated to seal both the supply and exhaust air from associated ductwork during the decontamination cycle. The CURIS^®^ decontamination unit was programmed, equipment positioned, and the room doors sealed using polyethylene sheeting and non-porous adhesive tape (Figure S2).

### Decontamination Process

The CURIS^®^ decontamination unit programming method utilizes room size to establish the baseline parameters for charge (initial aHP dispensing) and intermittent aHP pulse periods (additional aerosol pulses). This is followed by a user-defined dwell period (when no further aHP is introduced), at closure of the pulse period. Aeration to disperse residual H_2_O_2_ follows the dwell period (i.e. room seals are broken and ventilation resumed). Once the user inputs the room’s cubic volume or dimensions, the CURIS unit calculates a suggested duration of charge and pulse periods. The standard aeration period is 3 hours, unless an auxiliary scavenging system or other space aeration system is utilized. To account for absorption of aHP into porous materials in the decontamination space (e.g. N95 respirators), a 30-40% increase in these default settings was initially used, as recommended by the manufacturer. Additionally, to ensure adequate contact time of disinfectant to the treated surfaces, a dwell period was added to the continuous and pulse charge periods. Based on initial results, adjustments in the charge, pulse, and dwell periods were made to optimize the decontamination process (Tables 3-4).

After completion of the decontamination phase, an aeration or dissipation phase was initiated by removing fixed room seals and opening exhaust dampers for up to 2-hour periods, with the room under slight vacuum (0.18-0.19” W.G.). Adjustments were made to room air exchange rate during aeration to efficiently dissipate detectable H_2_O_2_ from respirator facepieces. At cycle 5, final parameters were established to include an aeration exhaust rate greater than or equal to 35 air changes/hour, with make-up air supplied by outdoor air. Following this phase, and after room H_2_O_2_ concentration was measured at less than 2 ppm, the respirator-holding rack was either retained under ventilation or transferred to a separate room (referred to as the “Finishing Room”) with an HVAC air supply curtain to further dry and decompose residual aHP from respirator facepieces to less than 1 ppm H_2_O_2_ (24, 25).

### Respirator Handling Process

For respirators subjected to repeated rounds of decontamination as part of this study, decontamination cycles were conducted repetitively from staging through drying. In order to be considered “dry” or ready for the next cycle, the interior and exterior respirator surfaces were monitored using a calibrated, hand-held real-time H_2_O_2_ monitor (ATI PortaSens II). Once H_2_O_2_ concentrations measured at respirator surfaces at less than 1 ppm, respirators were re-staged for the next round of decontamination, or packaged and transported for subsequent respirator fit-testing or virus inactivation analysis. Between each cycle, the treated/dried respirators were subjected to manual stress, by flexing each respirator bi-directionally and stretching each strap twice, using a hold position similar that used in respirator donning or doffing. A standard thin-line VWR Lab Marker was to mark each strap for each round of the aHP process (Figure S2C).

### Spore-based Biological Indicators

Commercial biological indicators (BI) were used for verification of decontamination. These commercially-prepared spore discs or “coupons” (Steris Spordex^®^) are enclosed in Tyvek/glassine envelopes (see Figure S3 for image) and contain a mean spore count of 2.4 x 10^5^ *Geobacillus stearothermophilus* (ATCC^®^ 7953;) (47). Between 6-12 BIs per cycle were placed throughout the room for each decontamination cycle. These were located behind or beneath equipment and surfaces, on the portable metal rack holding respirators, and either on or nested within the pairs of respirators to test aHP penetration (Figure S3). After each cycle, each BI spore disc was transferred from its glassine/Tyvek envelope to tryptic soy broth (Spordex^®^ Culture Media), incubated at 55°C, and analyzed after 7 days as an indicator of effective decontamination. There was one instance of a spore disc dropped during transfer, which resulted in a single positive BI from that cycle (cycle 5; see Table 4).

### Chemical Indicators of H2O2

Chemical indicator strips (Steris Steraffirm^®^ or CURIS^®^ System Hydrogen Peroxide Test Strips) (between 1 and 4 total per cycle) were placed in various locations throughout the Prep Room to indicate the presence of H_2_O_2_, supporting successful decontamination.

### Real-Time Hydrogen Peroxide Monitoring

The portable ATI PortaSens II detector (PortaSens) was used to measure H_2_O_2_ levels both within and outside the Prep Room during the decontamination process. Hydrogen peroxide concentrations were also measured at respirator surfaces, and necessarily reduced to less than 1 ppm prior to handling and sealing for transportation to designated tissue culture rooms, or for subsequent respirator fit-testing. During the charge and pulse periods of decontamination, real-time instantaneous sampling was conducted through a sealable wall port (designed for sterilizer tubing) into the Prep Room. The PortaSens was also used to monitor Prep Room concentrations at the start of the aeration phase, and to verify concentrations were reduced to less than 2 ppm for safe re-entry to the room (without respiratory protection); these measurements were made at breathing zone height (BZH) or 5 feet above floor level. H_2_O_2_ concentrations were also monitored outside the Prep Room door seal (see Figure S2), to check for any H_2_O_2_ leakage.

The U.S. Department of Labor/Occupational Safety and Health Administration (OSHA) and the American Conference of Governmental Industrial Hygienists (ACGIH) have established or adopted an eight-hour time-weighted average (TWA) occupational permissible exposure limit (PEL) to hydrogen peroxide of 1 ppm (54, 55). Though researchers and fit-test subjects were not anticipated to experience this eight-hour exposure level, 2 ppm H_2_O_2_ concentration was used as a safety threshold for room entry, and < 1 ppm for removal of respirators, sealing respirators for transport, and re-use by study participants. See Supplementary Text (and Table S2) for additional metrics used to verify that research personnel were not adversely exposed to H_2_O_2._

### Respirator Selection for the Study

The N95 respirator facepiece models examined in this study include a range of characteristics, including those identified as surgical N95 respirators (no exhalation valve to maintain sterile field), and non-surgical N95 respirators with an exhalation valve (see Table 1 and Figure S1). Respirator models were required to be successfully fitted by test subjects. Since fit test results varied widely by subject during early testing of Alpha ProTech respirators, this model was discontinued from further study. Fit-testing participants included experienced test subjects and/or administrators. Several respirator models were qualitatively fit-tested (QLFT) using Occupational Safety and Health Administration (OSHA) fit test protocols (saccharine challenge) described in the OSHA Respiratory Protection Standard (29 CFR 1910.134) (56). This included standard exercises to challenge respirator fit, over an approximate 8 minute, 30 second period.

Additionally, quantitative fit-testing (QNFT) was conducted using OSHA protocol requirements with a T.S.I, Inc. PortaCount Pro+ Model 8038 Fit Tester. This latter method employs condensation nucleus or particle counting technology (CNC or CPC) to measure aerosol concentration outside and inside the facepiece to determine a user fit-factor (57).

### Sample Size and Acceptance Criteria for Fit-Test Reliability

Study design for the QLFT endpoint was intended to rigorously evaluate user respirator to facepiece seal using the greatest-available pool of respirators (stockpiled model 3M 8511), while also providing representative feasibility data for the other respirator models (Table 1). The stockpiled 3M 8511 respirators were procured and collected by Penn State from 2006 to 2009.

The use of these respirators beyond the manufacturer’s recommended shelf-life enabled the study to proceed without consuming respirators that were more urgently needed by frontline workers, and it was supported by CDC crisis-capacity scenarios in force at the time (40, 41). All other respirator models (Table 1) used in this study were within their expiration date. All respirators new and unused at study initiation.

Acceptance criteria for 3M 8511 respirators required a minimum sample of 59 facepieces with no failures during QLFT in order to conclude with 95% confidence that at least 95% of 3M 8511 respirators maintain fit integrity after repeated use and decontamination. A total of seventy-seven 3M 8511 respirators were available for study purposes. A small number of respirators were allocated for QNFT and virology testing after QLFT, since these processes entailed respirator destruction (i.e. grommet insertion for QNFT or mask slicing for viral resuspension) and rendered them inaccessible for subsequent rounds of aHP.

### Respirator Fit-Testing

Prior to decontamination, respirators were labeled with a unique identifier. After decontamination, a hash mark was placed on the lower elastic band of each respirator to identify the round(s) of decontamination completed (Figure S2). Since the study was conducted during the COVID-19 pandemic, physical distancing and active clinical operating conditions limited the use of multiple test subjects. Therefore, two subjects were selected (one male, one female) to maximize varying size and facial features for fit-testing. Respirators were subjected to QLFT on the first, fifth, and tenth rounds of decontamination. A small number of respirators per model were allocated for QNFT during each round of fit testing; due to installation of metal grommet/probe for QNFT, these could not be reused for subsequent aHP cycles or fit testing. These were repurposed for subsequent virus inactivation testing in order to conserve overall respirator use. See Supplementary Text for additional details on QNFT metrics.

### NIOSH Filtration Efficiency Testing

To determine whether N95 respirators used in this study experienced filtration media breakdown as a result of sequential aHP disinfection cycles, several respirators were sent for independent laboratory analysis. These included eight 3M 8511 respirators subjected to cyclic aHP treatment and intermittent fit-testing, and two additional unused, untreated 3M 8511 respirators. Respirator filtration efficiency testing was performed by ICS Laboratories, Inc. (ICS) of Brunswick, Ohio. ICS is one of two firms in the U.S. authorized by the National Institute for Occupational Safety and Health (NIOSH) to perform respirator certification or re-certification.

An abbreviated “short-cycle” filtration efficiency verification test was conducted using the NIOSH Standard Test Procedure TEB-APR-STEP-0059 (58), which is summarized below. The standard test protocol includes initial respirator conditioning at 85 +/- 5% relative humidity (%RH) and 38 +/- 2°C for 25 hours prior to testing. This conditioning was intended to reflect active moisture load created by the respirator user. Following sealing of the respirator exhalation valve, and placing into the test instrument, full-load testing was performed. This testing included respirator challenge using 200 milligrams sodium chloride aerosol (mean count particle size distribution verified as 0.075 +/- 0.020 microns, with geometric standard deviation not exceeding 1.86). Sodium chloride aerosol was neutralized to a “Boltzman equilibrium state” (25 +/- 5°C, 30 +/- 10% relative humidity), and introduced at an airflow rate of 85 +/- 4 liters per minute, with periodic check and adjustment to maintain this flowrate. Instrumental analysis was conducted using a T.S.I Automated Filter Tester Model 8130A. Recorded data included flow rate, resistance, penetration, maximum penetration, and filtration efficiency.

The test protocol establishes the means for ensuring that the particulate filtering efficiency of N95 series filters used on non-powered respirators submitted for Approval, Extension of Approval, or examined during Certified Product Audits, meets the minimum certification standards set forth in 42 CFR, Part 84, Subpart K, §84.181.

### Virus Inoculation and Titration

Viruses used here included herpes simplex virus 1 (HSV-1) strain F, coxsackievirus B3 (CVB3), *Pseudomonas* phi6 bacteriophage (phi6) and SARS-CoV-2 isolate USA-WA1/2020. The key characteristics of these viruses are summarized in Table 2. All virus-inoculated materials were handled in accordance with the biosafety level (BSL) specified for that virus (see Table 2). Viruses were propagated in the same host cells as used for viral titration (see virus-specific sections below). In all aHP cycles, the respirators used for virus inactivation testing had been subjected to preceding total number of aHP cycles (see Table 4 for details). The input inoculum for each virus was set to the maximum available in each viral stock preparation.

For virus inactivation testing, respirator facepieces were inoculated with a controlled amount of one or more surrogate virus species (refer to Table 2 for complete list). Each virus was added in duplicate (or in quadruplicate during “Post 1-3” cycles of testing) droplets of 10 μL each, on one or more surfaces of each respirator type. The zone of viral inoculation was marked at the corners, to allow excision of the inoculated area after aHP treatment or air-drying. Droplets were allowed to air-dry or absorb fully onto each respirator inside a Class II biosafety cabinet (BSC), before proceeding further. Selected samples of each virus-inoculated respirator were left in ambient conditions, without aHP decontamination, as a “drying-only” virus control. The remainder of each virus-inoculated facepiece was subjected to aHP decontamination as described above (i.e. “decontaminated” samples). For each round of viral testing, the duration of air-drying was matched to the duration of time needed for aHP treatment of parallel respirators (i.e. including transport, respirator staging in the Prep Room, aHP decontamination, aeration, and return to the viral testing location). After aHP treatment, respirators, each virus-inoculated area was cut out of the “dried” or “decontaminated” respirators using dissection scissors. Each excised virus-spot encompassed all layers of the respirator, to capture any virus that had absorbed beyond the surface fabric.

### Herpes Simplex Virus 1 and Coxsackievirus B3 Quantification

Excised respirator areas inoculated with HSV-1 or CVB3 were transferred into individual Eppendorf tubes and resuspended in a 250 μL volume of cell media. Cell media consisted of Dulbecco’s Modified Eagle’s Medium (DMEM) supplemented with 10% fetal bovine serum (FBS) and penicillin-streptomycin (Pen/Strep; 100 U/mL; Thermo Fisher Scientific). The number of infectious units, or plaque-forming units (PFU), for these viruses was determined by limiting dilution onto confluent Vero detector cell monolayers (*Cercopithecus aethiops* monkey kidney cells, ATCC^®^ CCL-81™). Plaque formation was assessed at 72 hours post infection (hpi), after fixation and visualization of plaques using methylene blue staining. Duplicates of each virus-inoculated respirator piece were frozen for titration at a subsequent date, separate from the initial viral quantification. This allowed for experimental redundancy in terms of detector cell monolayers, and reduced handling time for the large numbers of virus-respirator inoculation sites.

### Bacteriophage phi6 Quantification

Excised respirator areas inoculated with phi6 bacteriophage were transferred into individual Eppendorf tubes and resuspended in 250 μL of King’s medium B (KB). These were then quantified on lawns of *Pseudomonas syringae pathovar phaseolicola* strain 1448A (*Pph*) using a previously described soft agar overlay protocol (59). Briefly, 100 µl logarithmic culture of *Pph* (OD_600_ ∼ 0.5) and 100 µl of the phage preparation were sequentially added to 3 ml of molten soft agar (0.7%) maintained at 55°C. The mixture was quickly poured on top of a KB agar plate and dried in the biosafety cabinet before transferring to a 28°C incubator. Alternatively, for enumeration of “inoculum” and “dried” samples, soft agar *Pph* lawns were prepared and 10 µl dilutions of phage preparation were spotted. Plaque forming units (PFU) were enumerated after 24-48 hours of incubation at 28°C.

### SARS-Coronavirus 2 Quantification

All experiments with SARS-CoV-2 were conducted in the Pell Laboratory under biosafety level 3 (BSL3) enhanced conditions using the USA-WA1/2020 isolate. SARS-CoV-2 was spotted onto masks in quadruplicate and treated with aHP or left in ambient BSL3 conditions for drying-only controls. Respirator areas inoculated with SARS-CoV-2 were then excised and resuspended in 250 µL of DMEM supplemented with sodium pyruvate, non-essential amino acids, antibiotics-antimycotics, and 2% FBS. After resuspension, viral titer was determined by tissue culture infectious dose 50 (TCID50) assay in 96-well plates, using Vero E6 cells. Briefly, 20 µL of resuspended sample was added to 4 wells containing 180 µL of resuspension media. The added samples were then serially diluted 10-fold down the plate, and the plates were incubated at 37 °C for 96-hours. At this time, cytopathic effect (CPE) was scored, and titer was calculated using the method of Reed and Muench (60).

## Data Availability

All data is included in this publication. Supplemental Table 1 is available from the authors & included in the peer-review submission.

## Acknowledgements

We appreciate the contributions of colleagues and scientists who provided intellectual input and/or materials for this work. These include Joyce Jose, Susan Hafenstein, Anthony Schmitt, Gregory Broussard, Michael Brignati, and Tim Simpson. Paul Turner and his lab graciously provided phi6 bacteriophage. We appreciate the contributions and support of Jim Crandall and the Penn State Environmental Health and Safety staff, as well Theresa Engle and other Occupational Medicine staff, who provided support in study implementation and fit-testing. We also thank Irene Miller and Pell Lab decontamination staff, researchers, clinical staff, as well as Neerav Goyal, Justin Soulier, and the healthcare providers of the Penn State Health Milton S. Hershey Medical Center and College of Medicine.

This work was a largely volunteer-based effort, with support from the Pennsylvania State University and The Penn State Institutes of Energy and the Environment. SARS-CoV-2 research in Dr. Troy Sutton’s laboratory was initiated by a seed grant from The Huck Institutes of Life Sciences at Pennsylvania State University, and Dr. Sutton’s research is further supported by the USDA National Institute of Food and Agriculture, Hatch project 4605.

## Supplementary Text

### Methods for hydrogen peroxide diffusion sampling (by H2O2 vapor monitors)

Diffusion samplers, also known as hydrogen peroxide vapor monitors (HPMs) were utilized to monitor long-duration H_2_O_2_ leakage outside the Prep Room entry (outside containment or OC), on research personnel during decontamination activities (breathing zone samples, clipped to lapel-collar), and within sealed respirator transport containers (prior to fit-testing) to verify that H_2_O_2_ concentrations remained less than 1 ppm throughout transport and fit-testing. Once the decontamination process was standardized with specific room ventilation parameters, ongoing personnel monitoring was discontinued.

HPM sampler analysis was conducted by Advanced Chemical Sensors, Inc. (ACS) of Longwood, FL utilizing an ACS HP-10 hydrogen peroxide vapor monitor, via modified OSHA Method VI-6 (colorimetric analysis). Analysis was subcontracted and completed by laboratories participating in the American Industrial Hygiene Association (AIHA) Laboratory Accreditation Program. The AIHA is an ISO/IEC 17025 accrediting body (61, 62).

### Additional methods on quantitative fit-testing metrics

Based on historical studies by the QNFT device and respirator manufacturers, N95 respirators have maximum breakthrough for particle sizes of 0.1 – 0.3 micron. Particles smaller or larger than this size range have increased filtration efficiency (greater than 95%). QNFT fit testing of N95 respirators therefore focuses on measuring this maximum penetration size range, to ensure that detectable leakage was due to facepiece seal leakage. OSHA has established a minimum QNFT passing fit factor of 100 for half-face respirators (including N95s or filtering-facepiece respirators) (56). The QNFT instrument manufacturer has established a maximum quantifiable fit factor of 200, as limited by measurement reliability and particle counting factors. QNFT fit factors equal to or exceeding 200 are therefore reported as 200(+). The reported QNFT data were utilized to verify successful fit reported by QLFT.

### Results for hydrogen peroxide diffusion sampling (by H2O2 vapor monitors)

Diffusion samplers, also known as hydrogen peroxide vapor monitors (HPMs) were utilized to assess researcher exposure levels to H_2_O_2_, with reference to the OSHA permissible exposure limit (PEL) and ACGIH Threshold Limit Value (TLV^®^) of 1 ppm, as an eight-hour time-weighted average (TWA). HPMs were also collected to monitor H_2_O_2_ build-up within respirator containers prior to fit-testing (for test subject safety), and to verify that H_2_O_2_ concentrations remained low in areas occupied by researchers (outside containment, OC) during the decontamination process (Table S2). All personal breathing zone HPM sampler results were reported below sample detection limits (< 0.08 ppm, <0.1 ppm) for researchers conducting aHP processes (up to 3 hours). All results for HPMs placed within sealed transport containers were reported below sample detection limits (< 0.03 ppm), indicating no residual H_2_O_2_ present through transport and respirator fit-testing. All results for HPMs collected as OC samples were reported below sample detection limits (< 0.08 ppm, cycle 1b) and < 0.2 ppm (cycle 11).

## Supplementary Tables

**Table S1.** **(Excel): aHP Viral Testing Values**Excel file of data underlying Figures 2-4, and Table 4. These include the number of plaque- forming units (PFU) for phi6, HSV-1, and CVB3, as well as TCID50(log10)/mL values for SARS-CoV-2. As noted in Figures 2-4, for samples where no virus was detected, these numbers were replaced with non-zero values to allow their visualization on the log-scale plots in Figures 2-4. HSV-1, CVB3, and phi6 "no virus detected" values were set to 0.5 (less than 1 plaque detected), and SARS-CoV-2 values were set to 1 (less than the limit of detection of 1.2 TCID50(log10)/mL).

**Table S2:**
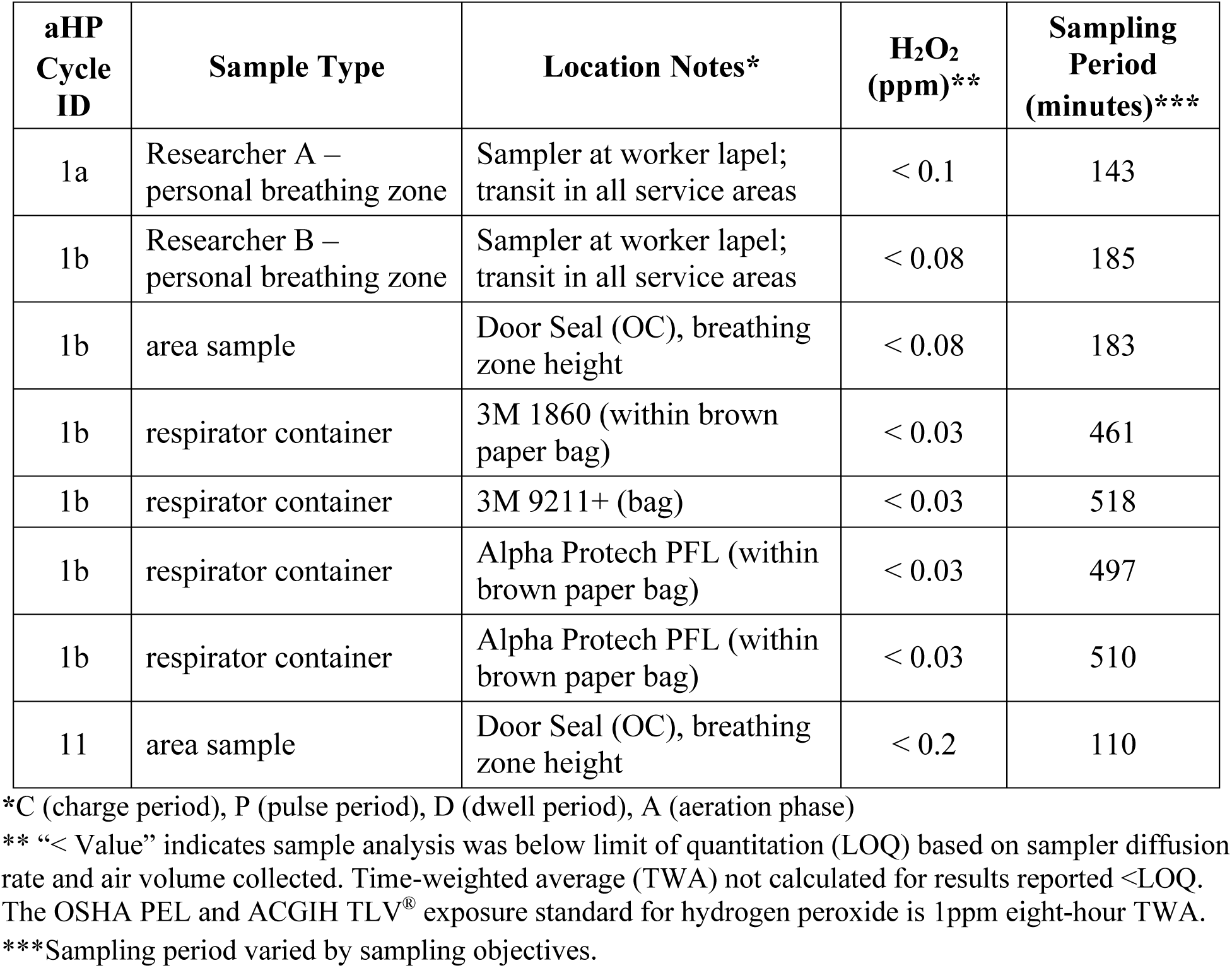
**Hydrogen peroxide diffusion sampler (HPM) data**

## Supplementary Figures

**Figure S1:**
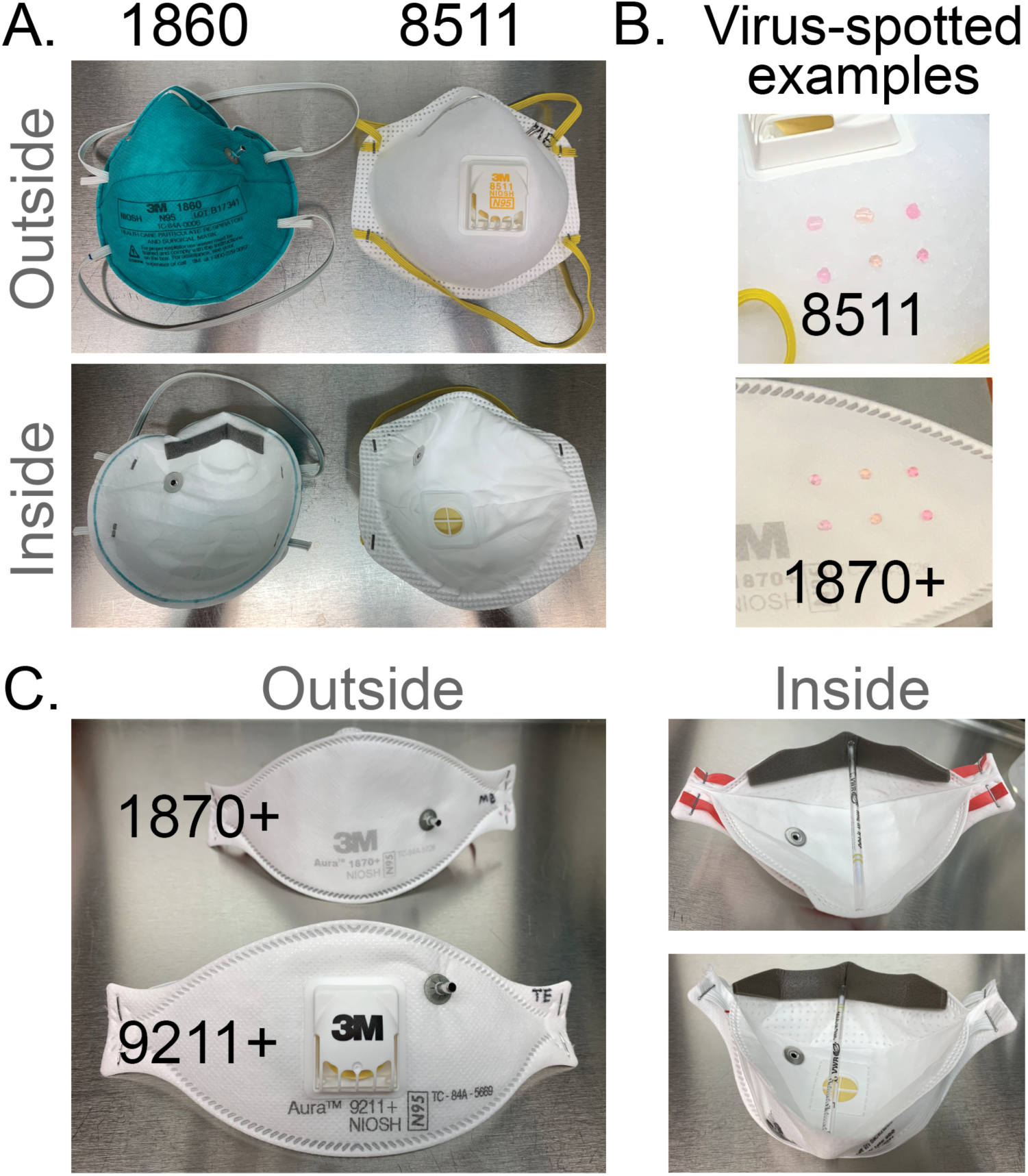
Respirator facepiece models & characteristics. (A) 3M models 1860 and 8511 (left) have a molded facepiece, with 8511 including an exhalation valve. (B) Virus-inoculation via droplets spotted onto the respirator fabric are shown for two examples. (C) 3M models 1870+ and 9211+ share a common folded fabric model, with a high fluid resistance of the fabric. 3M 9211+ includes an exhalation valve. Several of the facepieces (1860, 1870+, 9211+) display the metal grommet from insertion of the probe used for quantitative fit testing (QNFT). The inner face of each respirator is shown, which for the models shown in (C) required a prop to hold them open.

**Figure S2:**
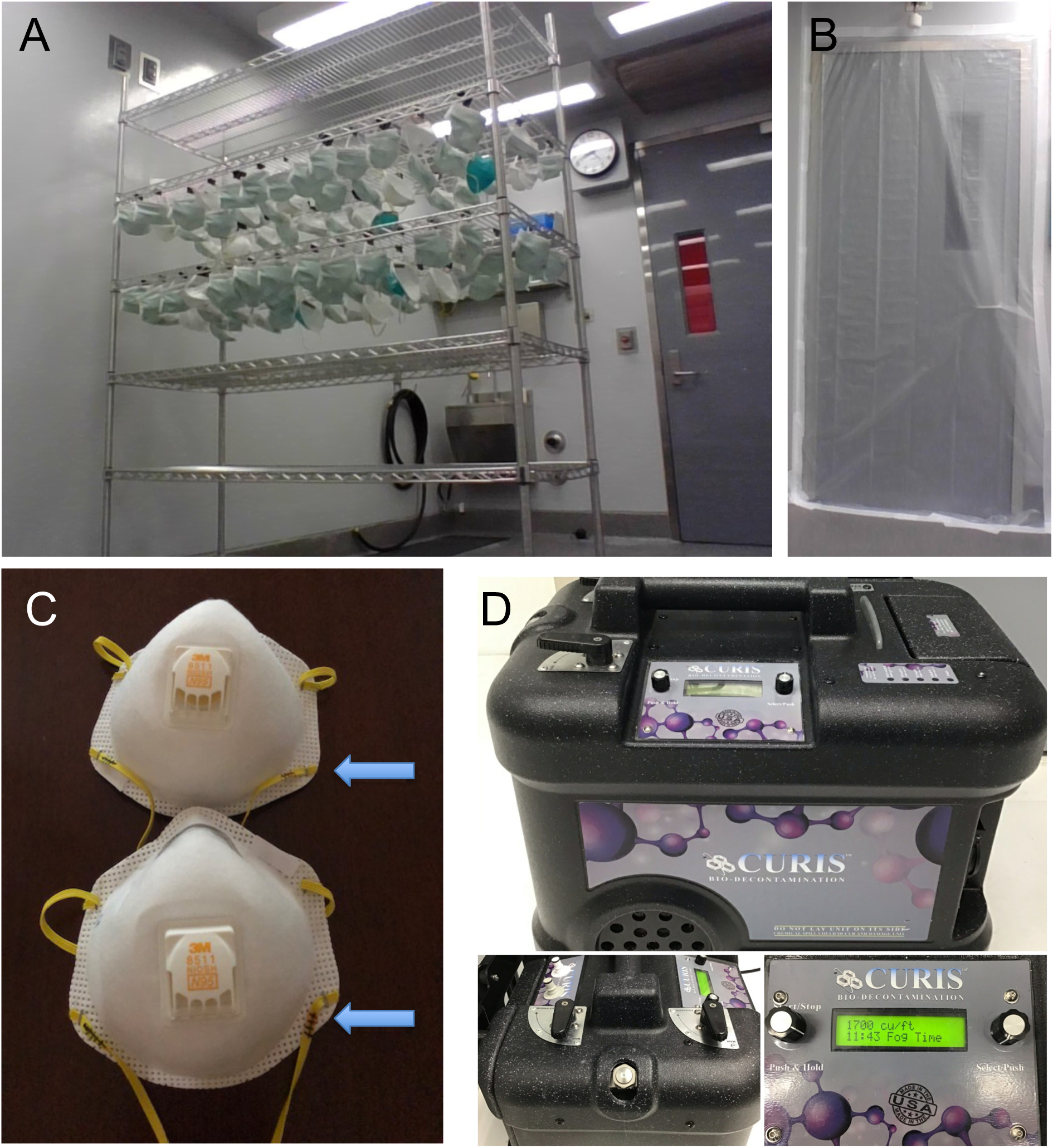
**Respirator staging and CURIS® unit.**(A) Respirator staging (partial loading) on metal rack with binder clips and rack centrally positioned in the Prep Room. (B) Prep Room door sealed with polyethylene sheeting and non- porous adhesive tape to prevent migration of H_2_O_2_ outside of the Prep Room. (C) Indelible ink pen hash marks (arrows) placed to notate completed aHP cycle, at time of cycle completion, or collection/transport for subsequent fit-testing or virus inactivation testing. (D) CURIS® decontamination unit with adjustable aHP dispersal nozzle and programmable inputs.

**Figure S3:**
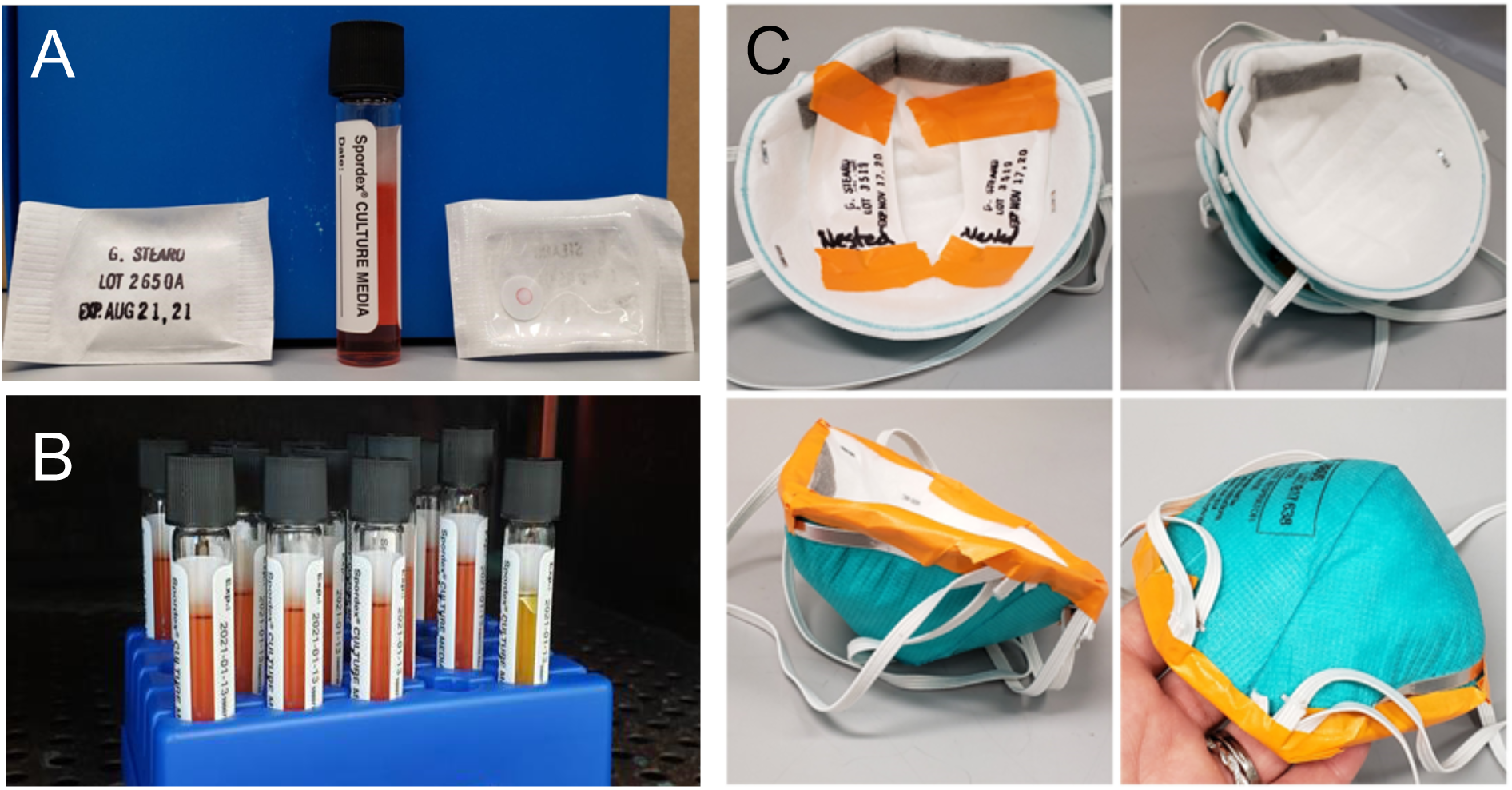
Spore-based biological indicators. (A) Commercial biological indicator (BIs; Steris Spordex^®^) containing *Geobacillius stearothermophilis* spores are sold on discs in Tyvek/glassine envelopes, with accompanying vials of culture media. After treatment, spore discs are transferred from to culture media and incubated for at 55°C for at least 7 days post decontamination cycle. (B) Media color change to yellow indicates bacterial growth, as seen in the positive control tube on the far right. In addition to placement of BIs around the treatment room (see Methods for details), BIs were nested between two molded respirators (3M 1860) and taped together (C), were enclosed within a flat-fold type respirator (3M Aura 9211+; not pictured). All BIs positioned in this manner were rendered inactivated after aHP treatment.

**Figure S4:**
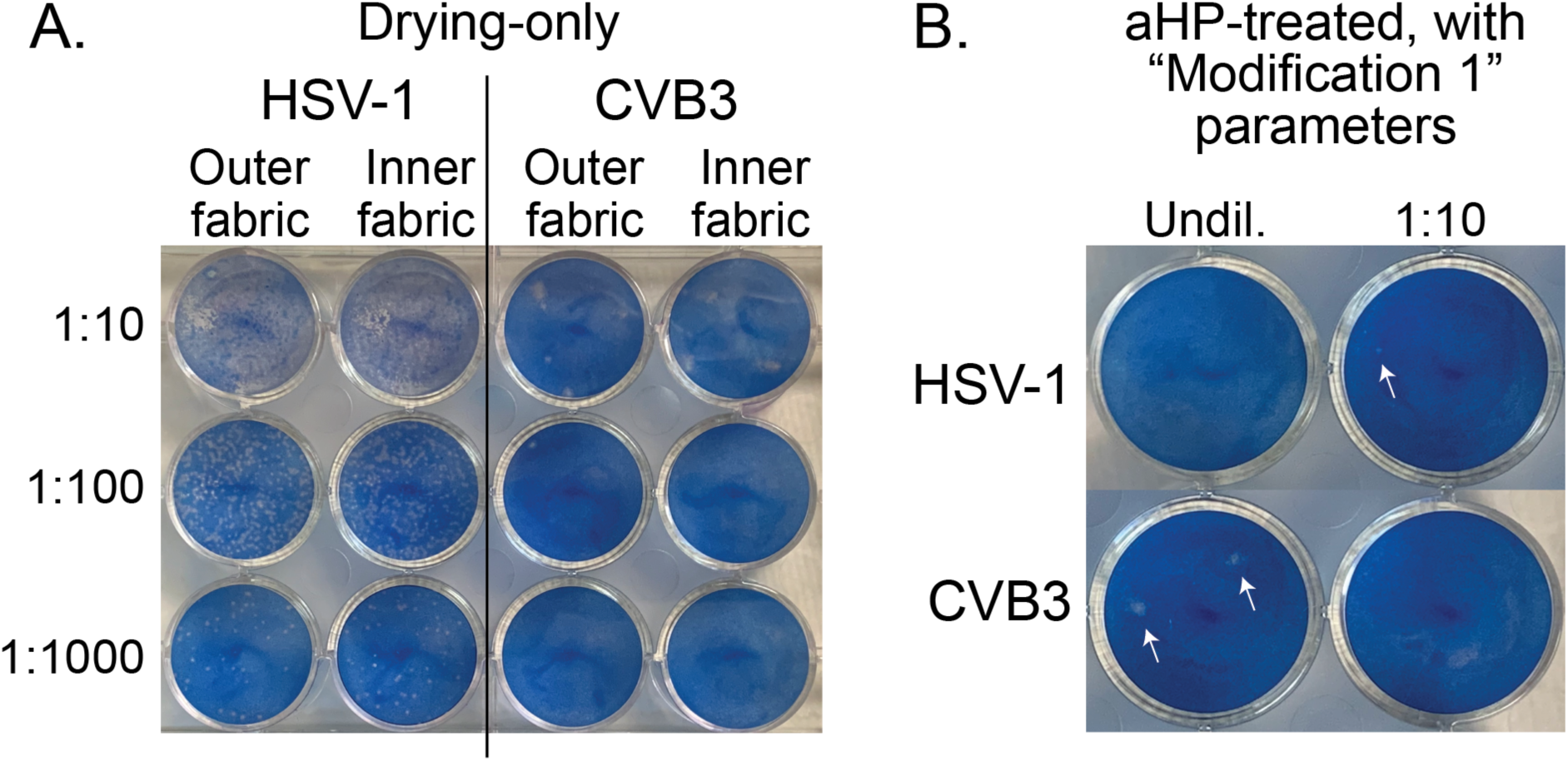
Viral titration demonstrates inactivation and loss of infectious units due to drying and aerosolized H_2_O_2_ (aHP) decontamination. Shown here are representative examples of serial dilutions (titration) of HSV-1 or CVB3 that were spotted onto 3M 9211+ respirators, and then resuspended and plated onto monolayers of Vero detector cells. Viral plaques, which are visible as clear foci of infection (plaque-forming units, or PFU) on the background of methylene blue-stained cells, were visualized at 72 hours post infection. The plate shown in **(A)** illustrates serial dilution of a high concentration of HSV-1 after drying-only (10^5^ PFU; see plot in Figure 3A), and a lower concentration of CVB3 after drying-only (10^2^ PFU; see plot in Figure 4A). The plate shown in **(B)** is from an aHP cycle run with “Modification 1” parameters, when no dwell time was used (aHP cycle 3; see Tables 3-4) and commercial spore-based biological indicators indicated a failure of decontamination. Even on this partial aHP decontamination, this sensitive detection method revealed only three wells (of which two are shown above) with any viral plaques (indicated by arrowheads; these equate to 25 PFU of HSV-1, and 5 PFU of CVB3; see plots in Figure 3A and 4A).

## Notes

### Competing Interest Statement

The authors have declared no competing interest.

### Summary of Updates

This manuscript includes revisions to address recommendations from peer review.

## References

1. Rutala WA, Weber DJ, Healthcare Infection Control Practices Advisory Committee (HICPAC). 2008. Guideline for Disinfection and Sterilization in Healthcare Facilities, 2008. Cent Dis Control CDC 163.

2. Centers for Disease Control and Prevention (CDC), National Center for Emerging and Zoonotic Infectious Diseases (NCEZID), Division of Healthcare Quality Promotion (DHQP). 2008. Guideline for Disinfection and Sterilization in Healthcare Facilities. Sterilizing Pract. https://www.cdc.gov/infectioncontrol/guidelines/disinfection/sterilization/sterilizing-practices.html.

3. 3M Personal Safety Division. 2020. Disinfection of Filtering Facepiece Respirators. 3M Tech Bull 4.

4. Battelle Memorial Institute. 2016. Final Report for the Bioquell Hydrogen Peroxide Vapor (HPV) Decontamination for Reuse of N95 Respirators. Prep Contract – Study Number 3245 US Food Drug Adm 46.

5. Liao L, Xiao W, Zhao M, Yu X, Wang H, Wang Q, Chu S, Cui Y. 2020. Can N95 Respirators Be Reused after Disinfection? How Many Times? ACS Nano 14:6348–6356.

6. Kumar A, Kasloff SB, Leung A, Cutts T, Strong JE, Hills K, Gu FX, Chen P, Vazquez- Grande G, Rush B, Lother S, Malo K, Zarychanski R, Krishnan J. 2020. Decontamination of N95 masks for re-use employing 7 widely available sterilization methods. PLOS ONE 15:e0243965.

7. Smith JS, Hanseler H, Welle J, Rattray R, Campbell M, Brotherton T, Moudgil T, Pack TF, Wegmann K, Jensen S, Jin J, Bifulco CB, Prahl SA, Fox BA, Stucky NL. 2020. Effect of various decontamination procedures on disposable N95 mask integrity and SARS-CoV-2 infectivity. J Clin Transl Sci 1–5.

8. Fischer RJ, Morris DH, van Doremalen N, Sarchette S, Matson MJ, Bushmaker T, Yinda CK, Seifert SN, Gamble A, Williamson BN, Judson SD, de Wit E, Lloyd-Smith JO, Munster VJ. 2020. Effectiveness of N95 Respirator Decontamination and Reuse against SARS-CoV-2 Virus. Emerg Infect Dis 26:2253–2255.

9. Schwartz A, Stiegel M, Greeson N, Vogel A, Thomann W, Brown M, Sempowski GD, Alderman TS, Condreay JP, Burch J, Wolfe C, Smith B, Lewis S. 2020. Decontamination and Reuse of N95 Respirators with Hydrogen Peroxide Vapor to Address Worldwide Personal Protective Equipment Shortages During the SARS-CoV-2 (COVID-19) Pandemic. Appl Biosaf 25:67–70.

10. Lieu A, Mah J, Zanichelli V, Exantus RC, Longtin Y. 2020. Impact of extended use and decontamination with vaporized hydrogen peroxide on N95 respirator fit. Am J Infect Control 48:1457–1461.

11. Cheng VCC, Wong S-C, Kwan GSW, Hui W-T, Yuen K-Y. 2020. Disinfection of N95 respirators by ionized hydrogen peroxide during pandemic coronavirus disease 2019 (COVID-19) due to SARS-CoV-2. J Hosp Infect 105:358–359.

12. Grillet AM, Nemer MB, Storch S, Sanchez AL, Piekos ES, Leonard J, Hurwitz I, Perkins DJ. 2020. COVID-19 global pandemic planning: Performance and electret charge of N95 respirators after recommended decontamination methods. Exp Biol Med 153537022097638.

13. Kenney PA, Chan BK, Kortright KE, Cintron M, Russi M, Epright J, Lee L, Balcezak TJ, Havill NL, Martinello RA. 2021. Hydrogen peroxide vapor decontamination of N95 respirators for reuse. Infect Control Hosp Epidemiol 1–3.

14. John AR, Raju S, Cadnum JL, Lee K, McClellan P, Akkus O, Miller SK, Jennings WD, Buehler JA, Li DF, Redmond SN, Braskie M, Hoyen CK, Donskey CJ. 2021. Scalable in-hospital decontamination of N95 filtering face-piece respirator with a peracetic acid room disinfection system. Infect Control Hosp Epidemiol 42:678–687.

15. Jatta M, Kiefer C, Patolia H, Pan J, Harb C, Marr LC, Baffoe-Bonnie A. 2021. N95 reprocessing by low temperature sterilization with 59% vaporized hydrogen peroxide during the 2020 COVID-19 pandemic. Am J Infect Control 49:8–14.

16. Levine C, Grady C, Block T, Hurley H, Russo R, Peixoto B, Frees A, Ruiz A, Alland D. 2021. Use, re-use or discard? Quantitatively defined variance in the functional integrity of N95 respirators following vaporized hydrogen peroxide decontamination during the COVID-19 pandemic. J Hosp Infect 107:50–56.

17. Russo R, Levine C, Grady C, Peixoto B, McCormick-Ell J, Block T, Gresko A, Delmas G, Chitale P, Frees A, Ruiz A, Alland D. 2021. Decontaminating N95 respirators during the COVID-19 pandemic: simple and practical approaches to increase decontamination capacity, speed, safety and ease of use. J Hosp Infect 109:52–57.

18. Christie-Holmes N, Tyli R, Budylowski P, Guvenc F, Weiner A, Poon B, Speck M, Naugler S, Rainville A, Ghalami A, McCaw S, Hayes S, Mubareka S, Gray-Owen SD, Rotstein OD, Kandel RA, Scott JA. 2021. Vapourized hydrogen peroxide decontamination in a hospital setting inactivates SARS-CoV-2 and HCoV-229E without compromising filtration efficiency of unexpired N95 respirators. Am J Infect Control 49:1227–1231.

19. Ludwig-Begall LF, Wielick C, Jolois O, Dams L, Razafimahefa RM, Nauwynck H, Demeuldre P-F, Napp A, Laperre J, Thiry E, Haubruge E. 2021. “Don, doff, discard” to “don, doff, decontaminate”—FFR and mask integrity and inactivation of a SARS-CoV-2 surrogate and a norovirus following multiple vaporised hydrogen peroxide-, ultraviolet germicidal irradiation-, and dry heat decontaminations. PLOS ONE 16:e0251872.

20. Laatikainen K, Mesilaakso M, Kulmala I, Mäkelä E, Ruutu P, Lyytikäinen O, Tella S, Humppi T, Salo S, Haataja T, Helminen K, Karppinen H, Kähkönen H, Vainiola T, Blomqvist K, Laitinen S, Peltonen K, Laaksonen M, Ristimäki T, Koivisto J. 2022. Large-scale decontamination of disposable FFP2 and FFP3 respirators by hydrogen peroxide vapour, Finland, April to June 2020. Eurosurveillance 27.

21. Oral E, Wannomae KK, Connolly RL, Gardecki JA, Leung HM, Muratoglu OK, Durkin J, Jones R, Collins C, Gjore J, Budzilowicz A, Jaber T. 2020. Vaporized H_2_O_2_ decontamination against surrogate viruses for the reuse of N95 respirators in the COVID-19 emergency. preprint. Occupational and Environmental Health.

22. Berger D, Gundermann G, Sinha A, Moroi M, Goyal N, Tsai A. 2021. Review of aerosolized hydrogen peroxide, vaporized hydrogen peroxide, and hydrogen peroxide gas plasma in the decontamination of filtering facepiece respirators. Am J Infect Control S0196655321004314.

23. Goyal SM, Chander Y, Yezli S, Otter JA. 2014. Evaluating the virucidal efficacy of hydrogen peroxide vapour. J Hosp Infect 86:255–259.

24. Tuladhar E, Terpstra P, Koopmans M, Duizer E. 2012. Virucidal efficacy of hydrogen peroxide vapour disinfection. J Hosp Infect 80:110–115.

25. Holmdahl T, Odenholt I, Riesbeck K, Medstrand P, Widell A. 2019. Hydrogen peroxide vapour treatment inactivates norovirus but has limited effect on post-treatment viral RNA levels. Infect Dis 51:197–205.

26. Hinton, Chief Scientist, Food and Drug Administration D. 2020. FDA Letter - Emergency Use Authorization (EUA) for the Battelle Critical Care Decontamination System (CCDS).

27. Hultman C, Hill A, McDonnell G. 2006. The physical chemistry of decontamination with gaseous hydrogen peroxide. Pharm Eng 27:22–32.

28. Otter JA, Yezli S. 2011. A call for clarity when discussing hydrogen peroxide vapour and aerosol systems. J Hosp Infect 77:83–84.

29. Boyce JM. 2009. New Approaches to Decontamination of Rooms After Patients Are Discharged. Infect Control Hosp Epidemiol 30:515–517.

30. Fu TY, Gent P, Kumar V. 2012. Efficacy, efficiency and safety aspects of hydrogen peroxide vapour and aerosolized hydrogen peroxide room disinfection systems. J Hosp Infect 80:199–205.

31. Holmdahl T, Lanbeck P, Wullt M, Walder MH. 2011. A Head-to-Head Comparison of Hydrogen Peroxide Vapor and Aerosol Room Decontamination Systems. Infect Control Hosp Epidemiol 32:831–836.

32. Freyssenet C, Karlen S. 2019. Plasma-Activated Aerosolized Hydrogen Peroxide (aHP) in Surface Inactivation Procedures. Appl Biosaf 24:10–19.

33. Best EL, Parnell P, Thirkell G, Verity P, Copland M, Else P, Denton M, Hobson RP, Wilcox MH. 2014. Effectiveness of deep cleaning followed by hydrogen peroxide decontamination during high Clostridium difficile infection incidence. J Hosp Infect 87:25–33.

34. Barbut F, Menuet D, Verachten M, Girou E. 2009. Comparison of the Efficacy of a Hydrogen Peroxide Dry-Mist Disinfection System and Sodium Hypochlorite Solution for Eradication of *Clostridium difficile* Spores. Infect Control Hosp Epidemiol 30:507–514.

35. Shapey S, Machin K, Levi K, Boswell TC. 2008. Activity of a dry mist hydrogen peroxide system against environmental Clostridium difficile contamination in elderly care wards. J Hosp Infect 70:136–141.

36. Bartels MD, Kristoffersen K, Slotsbjerg T, Rohde SM, Lundgren B, Westh H. 2008. Environmental meticillin-resistant Staphylococcus aureus (MRSA) disinfection using dry-mist-generated hydrogen peroxide. J Hosp Infect 70:35–41.

37. Andersen BM, Syversen G, Thoresen H, Rasch M, Hochlin K, Seljordslia B, Snevold I, Berg E. 2010. Failure of dry mist of hydrogen peroxide 5% to kill Mycobacterium tuberculosis. J Hosp Infect 76:80–83.

38. Beswick AJ, Farrant J, Makison C, Gawn J, Frost G, Crook B, Pride J. 2011. Comparison of Multiple Systems for Laboratory Whole Room Fumigation. Appl Biosaf 16:139–157.

39. Henneman JR, McQuade EA, Sullivan RR, Downard J, Thackrah A, Hislop M. 2022. Analysis of Range and Use of a Hybrid Hydrogen Peroxide System for Biosafety Level 3 and Animal Biosafety Level 3 Agriculture Laboratory Decontamination. Appl Biosaf 27:7– 14.

40. Centers for Disease Control and Prevention (CDC), National Center for Immunization and Respiratory Diseases (NCIRD), Division of Viral Diseases. 2020. Implementing Filtering Facepiece Respirator (FFR) Reuse, Including Reuse after Decontamination, When There Are Known Shortages of N95 Respirators. Coronavirus Dis 2019 COVID-19 Decontam Reuse Filter Facepiece Respir. https://www.cdc.gov/coronavirus/2019-ncov/hcp/ppe-strategy/decontamination-reuse-respirators.html.

41. Centers for Disease Control and Prevention. 2020. Strategies for Optimizing the Supply of N95 Respirators. Cent Dis Control Prev CDC. https://www.cdc.gov/coronavirus/2019-ncov/hcp/respirators-strategy/index.html. Retrieved 25 October 2021.

42. 3M Personal Safety Division. 2021. 3M^TM^ Aura^TM^ Health Care Particulate Respirator and Surgical Mask 1870+, N95. https://multimedia.3m.com/mws/media/890183O/3m-aura-healthcare-particulate-respirator-and-surgical-mask-1870-brochure.pdf. St. Paul, MN.

43. Rockey N, Arts PJ, Li L, Harrison KR, Langenfeld K, Fitzsimmons WJ, Lauring AS, Love NG, Kaye KS, Raskin L, Roberts WW, Hegarty B, Wigginton KR. 2020. Humidity and Deposition Solution Play a Critical Role in Virus Inactivation by Heat Treatment of N95 Respirators. mSphere 5.

44. Vasickova P, Pavlik I, Verani M, Carducci A. 2010. Issues Concerning Survival of Viruses on Surfaces. Food Environ Virol 2:24–34.

45. Kampf G, Todt D, Pfaender S, Steinmann E. 2020. Persistence of coronaviruses on inanimate surfaces and their inactivation with biocidal agents. J Hosp Infect 104:246–251.

46. Casanova LM, Jeon S, Rutala WA, Weber DJ, Sobsey MD. 2010. Effects of Air Temperature and Relative Humidity on Coronavirus Survival on Surfaces. Appl Environ Microbiol 76:2712–2717.

47. Steris Healthcare. Biological Indicators for Sterilization | STERIS. https://www.steris.com/healthcare/products/sterility-assurance-and-monitoring/biological-indicators. Retrieved 14 October 2020.

48. Firquet S, Beaujard S, Lobert P-E, Sané F, Caloone D, Izard D, Hober D. 2015. Survival of Enveloped and Non-Enveloped Viruses on Inanimate Surfaces. Microbes Environ 30:140– 144.

49. van Doremalen N, Bushmaker T, Morris DH, Holbrook MG, Gamble A, Williamson BN, Tamin A, Harcourt JL, Thornburg NJ, Gerber SI, Lloyd-Smith JO, de Wit E, Munster VJ. 2020. Aerosol and Surface Stability of SARS-CoV-2 as Compared with SARS-CoV-1. N Engl J Med https://doi.org/10.1056/NEJMc2004973.

50. Sizun J, Yu MWN, Talbot PJ. 2000. Survival of human coronaviruses 229E and OC43 in suspension and after drying onsurfaces: a possible source ofhospital-acquired infections. J Hosp Infect 46:55–60.

51. Duan S-M, Zhao X-S, Wen R-F, Huang J-J, Pi G-H, Zhang S-X, Han J, Bi S-L, Ruan L, Dong X-P, SARS Research Team. 2003. Stability of SARS coronavirus in human specimens and environment and its sensitivity to heating and UV irradiation. Biomed Environ Sci BES 16:246–255.

52. Rabenau HF, Cinatl J, Morgenstern B, Bauer G, Preiser W, Doerr HW. 2005. Stability and inactivation of SARS coronavirus. Med Microbiol Immunol (Berl) 194:1–6.

53. Geller C, Varbanov M, Duval R. 2012. Human Coronaviruses: Insights into Environmental Resistance and Its Influence on the Development of New Antiseptic Strategies. Viruses 4:3044–3068.

54. Occupational Safety and Health Administration (OSHA), United States Department of Labor. 2017. TABLE Z-1 Limits for Air Contaminants. 1910.1000 TABLE Z–1. https://www.osha.gov/laws-regs/regulations/standardnumber/1910/1910.1000TABLEZ1.

55. American Conference of Governmental Industrial Hygienists (ACGIH). 2019. Hydrogen Peroxide: Threshold Limit Values (TLV) Chemical Substances, 7th ed. Cincinnati, OH.

56. Occupational Safety and Health Administration (OSHA), United States Department of Labor. 2004. Fit Testing Procedures (Mandatory). 1910.134 App A. https://www.osha.gov/laws-regs/regulations/standardnumber/1910/1910.134AppA.

57. Operation and Service Manual. 2015. Portacount Pro 8030 and Portacount Pro+ 8038 Respirator Fit TestersRev. P. T.S.I., Inc., Shoreview, MN.

58. National Institute for Occupational Safety and Health. 2019. Determination of Particulate Filter Efficiency Level for N95 Series Filters Against Solid Particulates for Non-Powered, Air-Purifying Respirators Standard Testing Procedure (STP), p. 9. In TEB-APR-STEP- 0059Revision 3.2. https://www.cdc.gov/niosh/npptl/stps/apresp.html.

59. Hockett KL, Baltrus DA. 2017. Use of the Soft-agar Overlay Technique to Screen for Bacterially Produced Inhibitory Compounds. J Vis Exp https://doi.org/10.3791/55064.

60. Reed LJ, Muench H. 1938. A simple method of estimating fifty per cent endpoints. Am J Epidemiol 27:493–497.

61. Advanced Chemical Sensors. AIHA-LAP ID No. 102047. Longwood, Florida.

62. SGS Galson Laboratories. AIHA-LAP ID No. 100324. E. Syracuse, New York 13057.

